# Modeling the impact of Malakit intervention: one more step towards malaria elimination in the Guiana Shield?

**DOI:** 10.1101/2023.07.11.23292527

**Authors:** Yann Lambert, Raphaëlle Métras, Alice Sanna, Muriel Galindo, Helene Hiwat, Paola Marchesini, Stephen Vreden, Martha Cecilia Suárez-Mutis, Oscar Mesones Lapouble, Antoine Adenis, Mathieu Nacher, Pierre-Yves Boëlle, Chiara Poletto, Maylis Douine

**Affiliations:** Centre d’Investigation Clinique Antilles-Guyane (Inserm 1424), Centre Hospitalier de Cayenne Andrée Rosemon, Cayenne, French Guiana; Sorbonne Université, Inserm, Institut Pierre Louis d’Epidémiologie et de Santé Publique (IPLESP), Paris, France; Malaria program, Ministry of Health of Suriname, Paramaribo, Suriname; Malaria Technical Group, Vector Transmissible and Zoonotic Diseases Coordination, Ministry of Health, Brasilia, Brazil; Foundation for Scientific Research Suriname, Paramaribo, Suriname; Laboratory of Parasitic Diseases, Institute Oswaldo Cruz/Fiocruz, Rio de Janeiro, Brazil; Pan American Health Organization / World Health Organization (PAHO/WHO) Country Office in Suriname; Department of Molecular Medicine, University of Padova, Padova, Italy

**Keywords:** malaria elimination, hard-to-reach population, gold mining, French Guiana, impact evaluation, mathematical modeling

## Abstract

**Background:** Malaria elimination in mobile and hard-to-reach populations calls for new, tailored interventions. In the Guiana Shield countries, the malaria burden is high in the population working in illegal gold mining. Between April 2018 and March 2020, we implemented Malakit, a new intervention targeting gold miners, and relying on the distribution of kits for self-diagnosis and self-treatment. In this study, we evaluate the impact of Malakit on malaria transmission.

**Methods:** We fitted a mathematical model of malaria transmission to surveillance data from Brazil and Suriname, and to prevalence data from cross-sectional surveys, to estimate the change in treatment coverage and reproduction number between the pre-intervention (2014-2018) and intervention (2018-2020) periods.

**Results:** Model results show that treatment coverage of symptomatic all-species malaria infections increased from 26.4% (95%CrI 22.8, 30.3) prior intervention to 55.1% (95%CrI 49.9, 60.8) during the intervention, leading to a decrease of the reproduction number from 1.19 to 0.86. We estimate that on average 6943 all-species malaria infections were averted during the intervention, corresponding to a 48.7% reduction in incidence and 43.9% reduction in total infection prevalence.

**Discussion:** Malakit had a significant impact on malaria transmission by improving the access to treatment of the population working in illegal gold mining in French Guiana. Building on the regional efforts of the past twenty years, Malakit contributed to another step towards malaria elimination in the Guiana Shield.

## 1 Introduction

Addressing malaria in mobile, migrant and hard-to-reach populations remains a challenge for countries on the way to malaria elimination, and call for tailored and innovative strategies to improve access to diagnosis, treatment and vector protection (1–3). In the Amazon region, the communities involved in gold mining are a typical example of such populations with residual malaria (4). In French Guiana, the population working in illegal gold mining sites has been identified as the main reservoir of malaria in the region, predominantly infected with *Plasmodium falciparum*, and producing the majority of cases exported to the neighboring countries (5,6). Their lack of access to proper diagnosis and treatment translates into self-medication with under-the-counter antimalarials, a major risk for the emergence of artemisinin resistance.

To address this local and regional challenge, French Guiana, Suriname and Brazil jointly implemented a new intervention named Malakit. This intervention relied on the distribution of kits for self-diagnosis and self-treatment, associated to a training session, to gold miners going to work in French Guiana from the borders with Brazil and Suriname (7). Between April 2018 and March 2020, health facilitators trained 3733 participants and distributed 4766 kits, reaching about one third of the target population. The pre-post-evaluation of the intervention showed an improvement of knowledge and practices of gold miners towards malaria care and a drop in Polymerase Chain Reaction (PCR) prevalence of *Plasmodium* spp.. National malaria surveillance systems recorded a decrease of the incidence of symptomatic cases imported from French Guiana concomitant with Malakit implementation. A preliminary interrupted time series analysis (ITSA) estimated at 43% the reduction of cases notified during the implementation of Malakit (8); still, ITSA does not make explicit assumptions on the mechanisms involved in malaria transmission thus limiting the investigation of the effect of the Malakit intervention.

Here we developed a mechanistic mathematical model explicitly accounting for malaria transmission, case detection and Malakit uptake during the intervention period. We employed a Bayesian approach to fit the model to surveillance and cross-sectional surveys’ data to estimate changes in treatment coverage and reproductive ratio. Then, by comparing the model outputs from the pre-intervention period (2014-2018) with the intervention period (2018-2020) we quantified the impact of Malakit on malaria incidence and prevalence, considering *P. falciparum* (*Pf*), *P. vivax* (*Pv*) and both species combined.

## 2 Materials and Methods

### 2.1 Population and settings

French Guiana is a French overseas territory located in South America, sharing borders with Brazil in the East, and Suriname in the West. Ninety percent of its land (85 000 km^2^) is covered by rainforest with a soil rich in gold – a geological characteristic shared by the countries of Guiana Shield. This gold is the reason for the continuous presence of an estimated population of 10 000 men and women on 500 to 700 illegal gold mining sites (9). The majority of gold miners are born in Brazil (>95%), 75% are men, with a median age of 38 (IQR: 31, 47), and less than half have a secondary level of education (5,8).

### 2.2 Intervention

The Malakit intervention has been described in several articles (7,10,11). Coordinated by Cayenne Hospital, Malakit was implemented between April 2018 and March 2020 at five sites mainly located at the borders of French Guiana with Brazil and Suriname (Figure 1). These sites were selected because of their strategic importance as entry points for gold miners going to French Guiana. The core intervention relied on the distribution of free kits for the self-diagnosis and self-treatment of malaria to gold miners by health facilitators. One kit contained three non-species-specific rapid diagnostic tests (RDT) for malaria, a full course of artemisinin combined therapy (ACT) (artemether - lumefantrine) and a single dose of primaquine (S-PQ). Before distributing a kit, health facilitators trained the participants to use the kit by making them perform an RDT themselves and explaining how to take the medication. Long lasting impregnated nets (LLINs) were also distributed to participants during the full course of the intervention in Suriname, and during the last six months of the intervention in Brazil.

**Figure 1:**
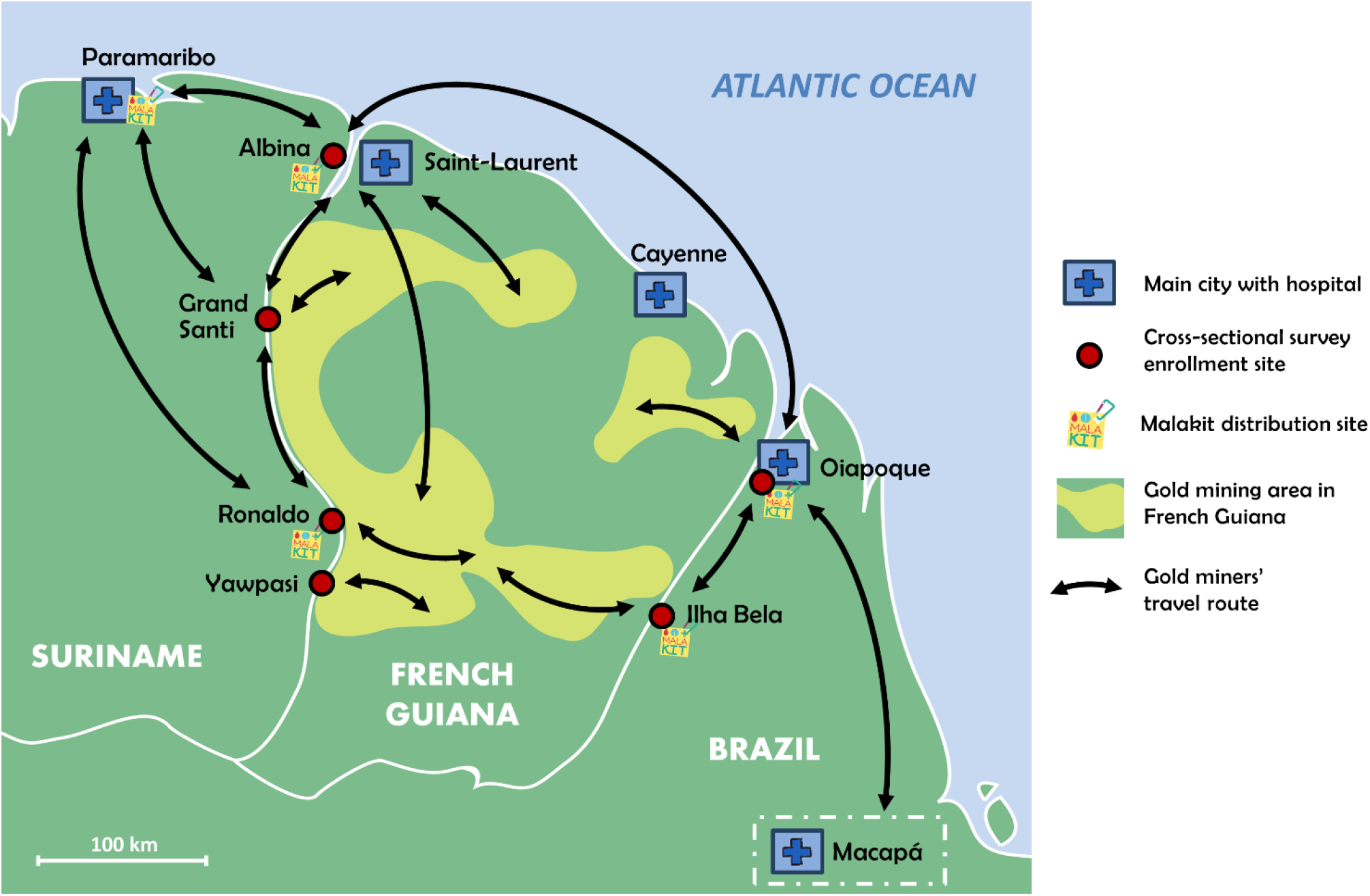
Map showing the location of Malakit distribution sites and pre/post intervention cross-sectional surveys enrolment sites at the borders of French Guiana with Brazil and Suriname

### 2.3 Malaria prevalence and surveillance data

We conducted cross-sectional surveys before and after the intervention to assess knowledge and self-reported practices about malaria treatment and diagnosis, and to estimate the prevalence of *Plasmodium* carriage by PCR in the population (5,8). Monthly notification data of malaria cases imported from French Guiana to Brazil and Suriname were obtained from national surveillance systems of the Brazilian and Surinamese malaria programs. From these surveillance data, we used only cases imported from French Guiana, and explicitly recorded as originating from gold mining. We chose not to use surveillance data from French Guiana because they do not allow consistently to identify cases specifically originating from mining areas.

### 2.4 Malaria transmission model

We developed a deterministic Susceptible-Infectious-Susceptible (SIS) compartmental model to simulate the transmission of malaria in a single population encompassing all individuals living in illegal gold mining sites in French Guiana. We used a human-to-human transmission rate which accounted for the vector population dynamics (12–14) with a seasonal forcing defined as an exponential effect of maximum daily temperature, with a lag. In our model, the infected population (I) corresponded to individuals with a detectable parasite density in PCR and was distributed into four model compartments: asymptomatic (IA), symptomatic but not (or incompletely) treated (IS) and symptomatic treated either by the health system (IT) or with the use of a malakit (IM). Susceptible individuals (S) were free from disease. We assumed that cases notified to the malaria surveillance systems (D) was a fraction of the infected individuals receiving treatment from health systems (IT) (Figure 2).

**Figure 2:**
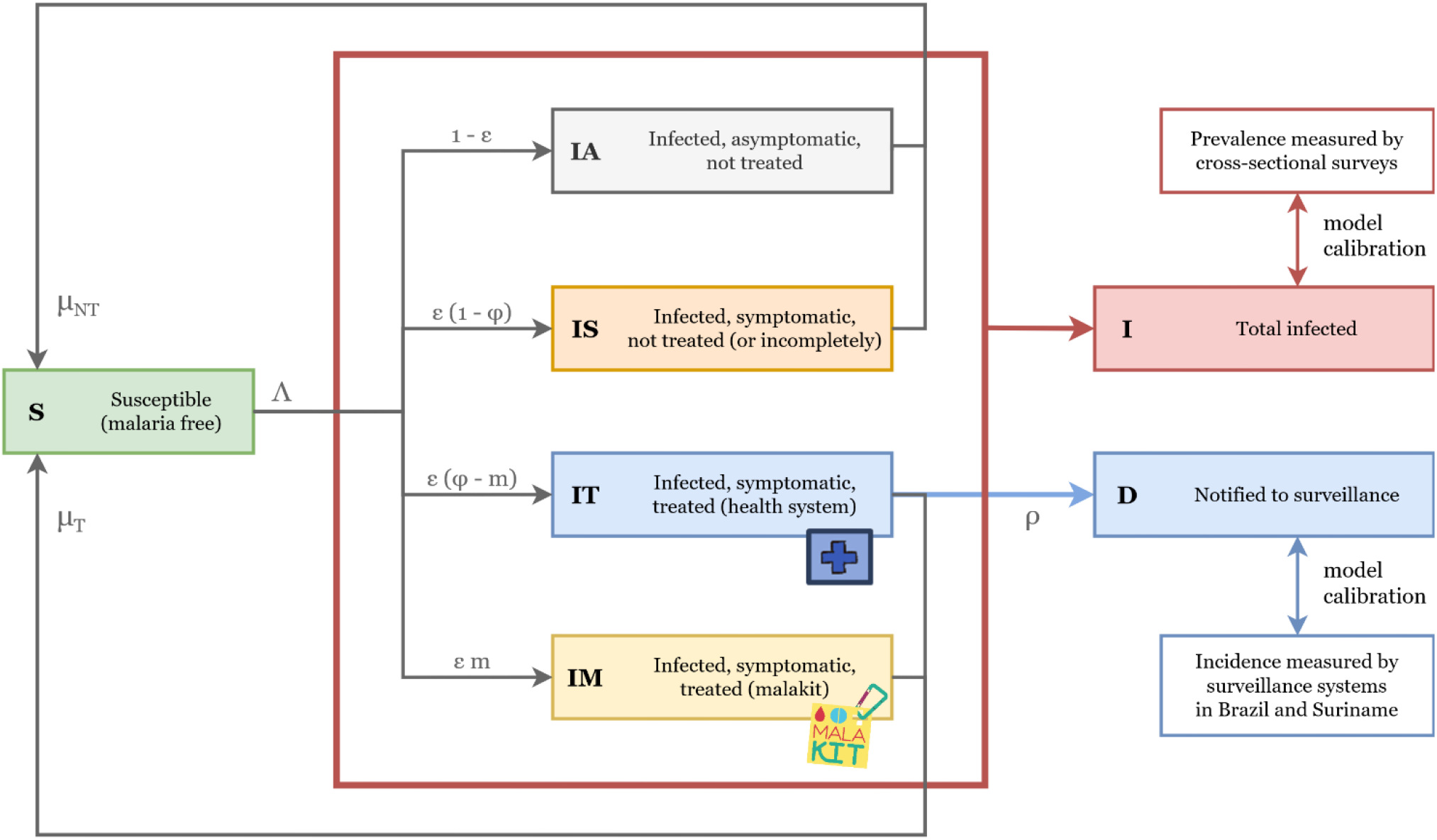
Compartments used in the malaria transmission model and sources of data used for calibration. Susceptible individuals (S) may get infected with malaria without symptoms (IA), or develop symptoms without appropriate treatment (IS), get treatment from the health system (IT) or from using a malakit (IM, during intervention). A fraction of the cases treated by the health system are notified to malaria surveillance (D).

Our model relied on the following assumptions: (i) the population size is constant, (ii) the mixing of individuals is homogeneous, (iii) the force of infection depends on meteorological conditions, with temperature chosen as explanatory variable; (iv) infected individuals have a constant probability *ε* of being symptomatic; (v) the rate of transmission from asymptomatic individuals is lower than from symptomatic individuals, with a constant ratio *k* < 1; (vi) only symptomatic individuals are treated with probabilities φ_0_ and φ_1_, prior and during Malakit implementation respectively; (vii) prior to Malakit implementation all individuals are treated by the health systems (φ_0_), while during the intervention period a proportion *m* of symptomatic individuals is treated with Malakit and a proportion φ_1_ − *m* is treated by health systems; (viii) the proportion of individuals who seek care from the health remains the same during the intervention period (φ_0_ = φ_1_ − *m*); (ix) treatment shortens the infection duration; (x) treatment does not affect the transmission rate; (xi) without treatment, the duration of infection is the same for asymptomatic and symptomatic individuals; (xii) individuals who seek treatment from the health system are notified to malaria surveillance in Brazil and Suriname with a constant reporting fraction *ρ*; and (xiii) individuals using Malakit for treatment are not reported to malaria surveillance.

Model equations and parameters definitions are presented in the supplementary methods.

### 2.5 Parameters estimation

We fitted our model to prevalence and surveillance data for 1) all species, 2) *Pf*, and 3) *Pv*. We estimated the posterior distribution of six parameters: probability of treatment before intervention (φ_0_), probability of treatment during intervention (φ_1_), probability of symptomatic infection (*ε*), transmission rate at average maximum daily temperature (*β*_0_), temperature-associated amplitude of transmission rate (*α*) and temperature-associated lag of the transmission rate (*lag*). We fixed the values of the reporting fraction (*ρ*=0.69), the duration of infection with treatment (*T*_*T*_=7 days), the initial number of infected individuals (*I*_0_, specie-specific), the duration of infection without treatment (*T*_*NT*_, specie-specific), and the ratio of asymptomatic vs. symptomatic transmission rates (*k*, specie-specific). Reporting fraction was *ρ*=0.69, while the duration of infection with treatment was *T*_*T*_=7 days. Values of *I*_0_, *T*_*NT*_, and *k* for *Pf, Pv* or both species combined are reported in the supplementary materials.

The log-likelihood associated to parameters estimates was calculated by fitting monthly simulated notifications to surveillance data assuming Poisson distribution, and simulated prevalence to 2015, 2018 and 2019 data assuming binomial distribution.

We used R (v4.2.0) and RStudio (v2022.02.3) to run three Markov chains of 100 000 iterations using different seed values, for each model fit. We used the Robust Adaptative Metropolis algorithm developed by Vihola (15) and implemented in R by Helske (16). We discarded the first 20 000 iterations of the burn-in periods and thinned at a ratio of 1:10 to eliminate auto-correlation and combined the three chains. We checked for convergence to the same stationary distribution both visually and using Gelman statistics from coda R package (17) (supplementary Table 10).

At each iteration of the Markov chains, we computed the probability of using a malakit, *m*, from fitted values of φ_0_ and φ_1_; the mean infection duration (*T*) and the reproduction number (*R*), before and during intervention, and derived the mean and 95%CrI from the resulting distributions. We performed a sensitivity analysis of the variation of the parameters’ posterior distribution for different values of *I*_0_ and *T*_*NT*_.

### 2.6 Estimation of the impact of Malakit

To assess the impact of the Malakit intervention on malaria incidence and prevalence, we ran simulations parameterized from a sample of 1000 draws from the parameter posterior distributions. For each one of the 1000 draws of parameter values, we compared the model simulation to a matching counterfactual (without intervention) with φ_1_ set to the pre-intervention value φ_0_. We then calculated the difference between the mean prevalence, the mean annual incidence rate and the cumulated incidence estimated by each pair of simulations, and derived the mean and 95%CrI from the resulting distributions.

### 2.7 Ethics and regulation

Ethics approvals the were obtained from countries where the Malakit study was implemented (8). All data from the Malakit project (intervention and cross-sectional surveys) were obtained anonymously after a written consent from participants. The analysis of all data complies with the General Data Protection Regulation (European Union Regulation 2016/679).

## 3 Results

### 3.1 Impact on all-species malaria transmission

Table 1 presents the values of estimated parameters. Over the full period 2014-2020 we estimate that 47.1% (95%CrI 38.8, 56.6) of new all-species malaria infections were symptomatic. Our model estimates the treatment coverage of symptomatic infections at 26.4% (95%CrI 22.8, 30.3) prior to the implementation of the Malakit intervention and 55.1% (95%CrI 49.9, 60.8) during the intervention, corresponding to a two-fold increase. According to our assumptions, this improvement of treatment coverage corresponds to a probability of 28.7% (95%CrI 25.0, 32.6) of using Malakit to treat a new symptomatic infection (NSI). This evolution translates into a 12-day decrease of the mean duration of infection (from 45 to 33 days, asymptomatic and symptomatic infections combined) and thus to a reduction of all-species malaria transmission, with a reproduction number (at average temperature) estimated at 1.19 (95%CrI 1.16, 1.22) prior intervention and 0.86 (95%CrI 0.83, 0.90) during the intervention.

**Table 1:** Posterior distributions (mean and 95% credible interval CrI) for all-species malaria transmission model.

| Parameter | Pre-intervention, 2014-2018<br>(95%CrI) | Intervention, 2018-2020<br>(95%CrI) |
| --- | --- | --- |
| $\phi$: Treatment coverage | 26.4% (22.8, 30.3) | 55.1% (49.9, 60.8) |
| $m$: Probability of using a kit in case of symptoms* | - | 28.7% (25.0, 32.6) |
| $\epsilon$: Probability of symptoms | 47.1% (38.8, 56.6) | |
| $\beta_0$: Transmission rate at average temperature ($\text{day}^{-1}$) | 0.027 (0.023, 0.031) | |
| $\alpha$: Seasonal amplitude of transmission rate ($^{\circ}\text{C}^{-1}$) | 0.097 (0.078, 0.115) | |
| lag: Temperature lag of transmission rate (days) | 83 (73, 94) |  |
| T: Mean infection duration (all infections) (days)* | 45 (39, 52) | 33 (28, 38) |
| R: Mean reproduction number (all infections)* | 1.19 (1.16, 1.22) | 0.86 (0.83, 0.90) |

\* Calculated from other parameters (see supplementary methods)

In the sensitivity analysis conducted for different values of T_NT_ and I_0_, most parameters showed little variation aside from ε. Importantly, computed values of reproductive number, prior and during intervention, showed little or no variation. The mean estimates of φ_0_ and φ_1_ increased from 0.22 to 0.28, and from 0.50 to 0.57, respectively, however with consistently overlapping 95%CrIs (supplementary Table 7).

Figure 3 shows the model fit to surveillance and prevalence data for all-species cases of malaria. The mean annual incidence rate dropped from 366 NSI/1000/year (95%CrI 304, 428) to 169 (95%CrI 131, 207), prior vs. during the intervention. In parallel, the mean notification rate is estimated to have decreased from 66 NSI/1000/year (95%CrI 50, 82) prior intervention, to 31 (95%CrI 20, 42) during intervention, representing a reporting fraction of 18.2%. Without intervention, the counterfactual scenario predicts that the mean incidence rate could reach 330 NSI/1000/year (95%CrI 262, 398). Therefore, we estimate that Malakit contributed to a 48.7% reduction of the all-species malaria incidence (intervention vs. counterfactual) and accounted for 81.5% of the total decrease of incidence since its implementation.

**Figure 3:**
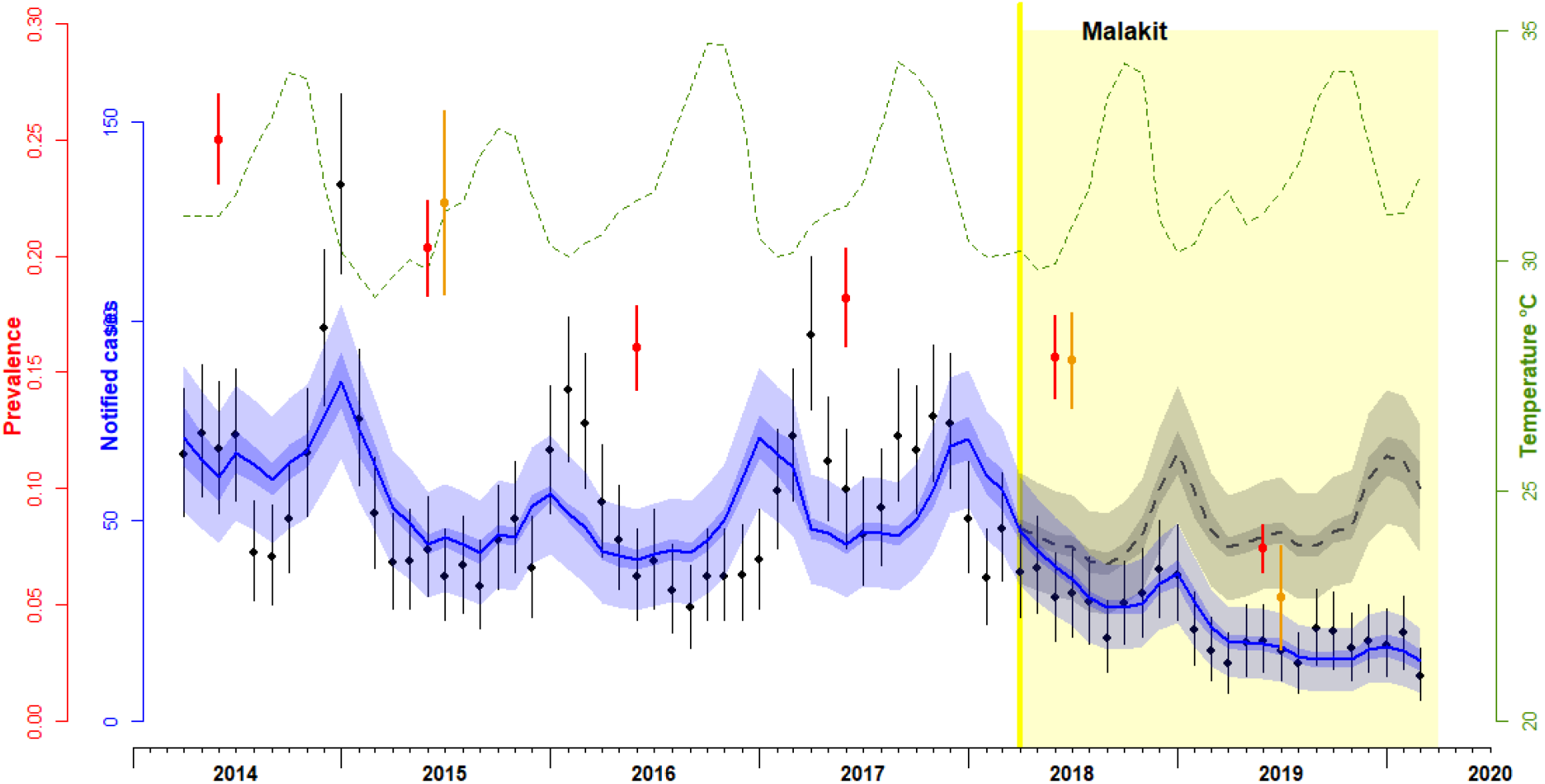
Monthly incidence of all-species reported cases of malaria (black dots), model-fitted simulations (blue solid line) and counterfactual estimates (dashed grey line). Annual all-species prevalence measured in PCR surveys (orange dots) and model-fitted estimates of prevalence (red dots). Maximum daily temperature (monthly average) is shown in green dashed line. The period of implementation of Malakit is shown in yellow. Vertical lines represent 95%CrI. For model-fitted and counterfactual incidence (respectively blue and grey), the dark envelopes show the uncertainty (95%CrI) associated to the joint posterior distribution, while the light envelopes account also for the uncertainty resulting from of the observed Poisson process.

The total prevalence (symptomatic and asymptomatic infections) is estimated to have decreased on average from 19.6% (95%CrI 17.8, 21.4) before intervention to 9.6% (95%CrI 8.5, 10.7) during intervention. Without intervention, counterfactual simulations predict that the prevalence could decrease to 17.1%. We thus estimate that Malakit led to a reduction of 43.9% of all-species malaria prevalence (supplementary Table 6).

Without intervention, the model predicts that the burden of all-species malaria would have reached a total of 14 053 symptomatic and asymptomatic infections between 2018 and 2020 (Table 2). With the Malakit intervention, this burden is estimated at 7205 new infections overall. We thus estimate that 6848 infections were averted during the two years of the intervention, of which 3218 symptomatic infections. According to our assumptions, we estimate that 970 infections were treated with the use of Malakit.

**Table 2:** Cumulated malaria incidence of all-species malaria infections estimated between April 2018 and March 2020, with intervention (model fitted to data) and without (counterfactual) (mean and 95%CrI).

|  | 2018-2020 cumulated incidence, all-species malaria infections (95%CrI) |  |  |  |
| --- | --- | --- | --- | --- |
|  | No intervention | Intervention | Difference | Variation |
| Notified (D) | 1193 (1101, 1286) | 612 (548, 676) | 581 (469, 694) | -48.7% (-55.1, -41.7) |
| Symptomatic, treated by the health system (IT) | 1730 (1607, 1852) | 887 (803, 971) | 843 (694, 991) | -48.7% (-54.5, -42.4) |
| Symptomatic, treated with a malakit (IM) | - | 970 (778, 1163) | - | - |
| **Total symptomatic treated (IT+IM)** | **1730 (1607, 1852)** | **1857 (1647, 2067)** | **-127 (-371, 116)** | **+7.4% (-6.4, 22.3)** |
| Symptomatic, not treated (IS) | 4870 (3771, 5964) | 1525 (1103, 1947) | 3345 (2166, 4518) | -68.7% (-78.6, -55.4) |
| **Total symptomatic (IT+IM+IS)** | **6600 (5436, 7761)** | **3382 (2806, 3959)** | **3218 (1919, 4514)** | **-48.7% (-60.0, -34.2)** |
| Asymptomatic, not treated (IA) | 7453 (5765, 9138) | 3823 (2926, 4717) | 3630 (1723, 5537) | -48.7% (-63.3, -28.6) |
| **Total infected (I)** | **14 053 (12 550, 15 559)** | **7205 (6408, 8002)** | **6848 (5147, 8552)** | **-48.7% (-56.1, -40.2)** |

### 3.2 Impact on *P. falciparum* and *P. vivax* malaria transmission

When fitted separately to *Pf* or *Pv* data, the model estimates a stronger increase in treatment coverage for *Pf* (from 20.6% to 81.7%) than *Pv* (from 20.8% to 37.2%) (Table 3). The probability of using Malakit is higher for *Pf* (61.1%) than *Pv* (16.4%). This is associated to a sharper decrease of R for *Pf* than *Pv*, 0.96 to 0.34 (−64.2%) vs. 1.13 to 0.95 (−15.9%), respectively.

**Table 3:**
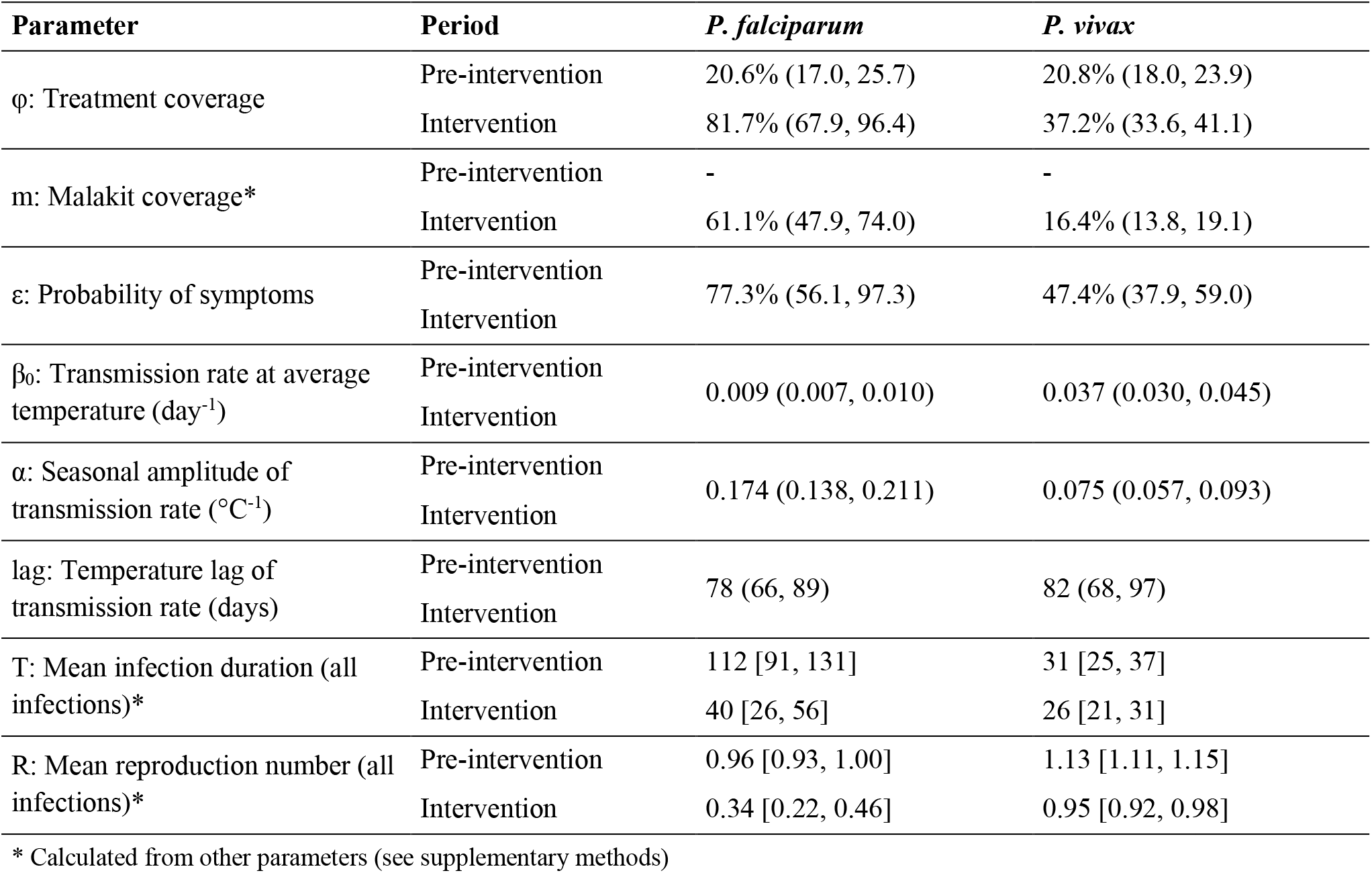
Posterior distributions (mean and 95% credible interval) for *P. falciparum* and *P. vivax* malaria transmission models.

We find species-specific patterns of malaria transmission: the probability of developing a symptomatic infection (ε) is higher for *Pf* than *Pv*: 77.3% vs. 47.4%. The transmission rate of *Pv* is about four times higher than *Pf* (mean *β*_0_ estimates: 0.037 vs. 0.009 day^-1^), however the seasonality of transmission is more marked for *Pf* than *Pv*, with seasonal amplitudes of transmission rate (α) estimated at 0.17 °C^-1^ vs. 0.08 °C^-1^, respectively.

When comparing intervention to counterfactual estimates, the estimated impact of Malakit on incidence rate is stronger for *Pf* (−63.1%) than *Pv* (−42.8%) (supplementary Table 6). However, the estimated contribution of Malakit to the total variation of the incidence rate since the beginning of the intervention is more modest for *Pf* (52.0%) than *Pv* (88.6%). A similar pattern is observed with the total prevalence for both species. These findings reflect the stronger decrease of *Pf* infection incidence and prevalence compared to *Pv* for reasons other than Malakit, in line with a mean reproduction number estimated lower than one before intervention for *Pf*, but not for *Pv*.

We estimate that 713 (95%CrI 507, 920) new *Pf* and 3686 (95%CrI 3067, 4303) new *Pv* symptomatic infections occurred during the intervention period (Table 4). Without intervention, the counterfactual scenario predicts that this burden would have reached 1944 (95%CrI 1336, 2550) *Pf* and 6452 (95%CrI 5311, 7591) *Pv* infections. We thus estimate that between 2018 and 2020 Malakit helped avert a total of 1231 NSI of *Pf* (95%CrI 589, 1870) and 2766 NSI of *Pv* (95%CrI 1471, 4063). The model estimates that 432 (95%CrI 311, 554) *Pf* and 605 (95%CrI 468, 742) *Pv* infections were treated by a malakit during the intervention. In line with a stronger increase of treatment coverage for *Pf* than *Pv*, we find a larger decrease of the incidence of symptomatic infections not treated (or incompletely) for *Pf* (−91.3%) than *Pv* (−54.6%).

**Table 4:**
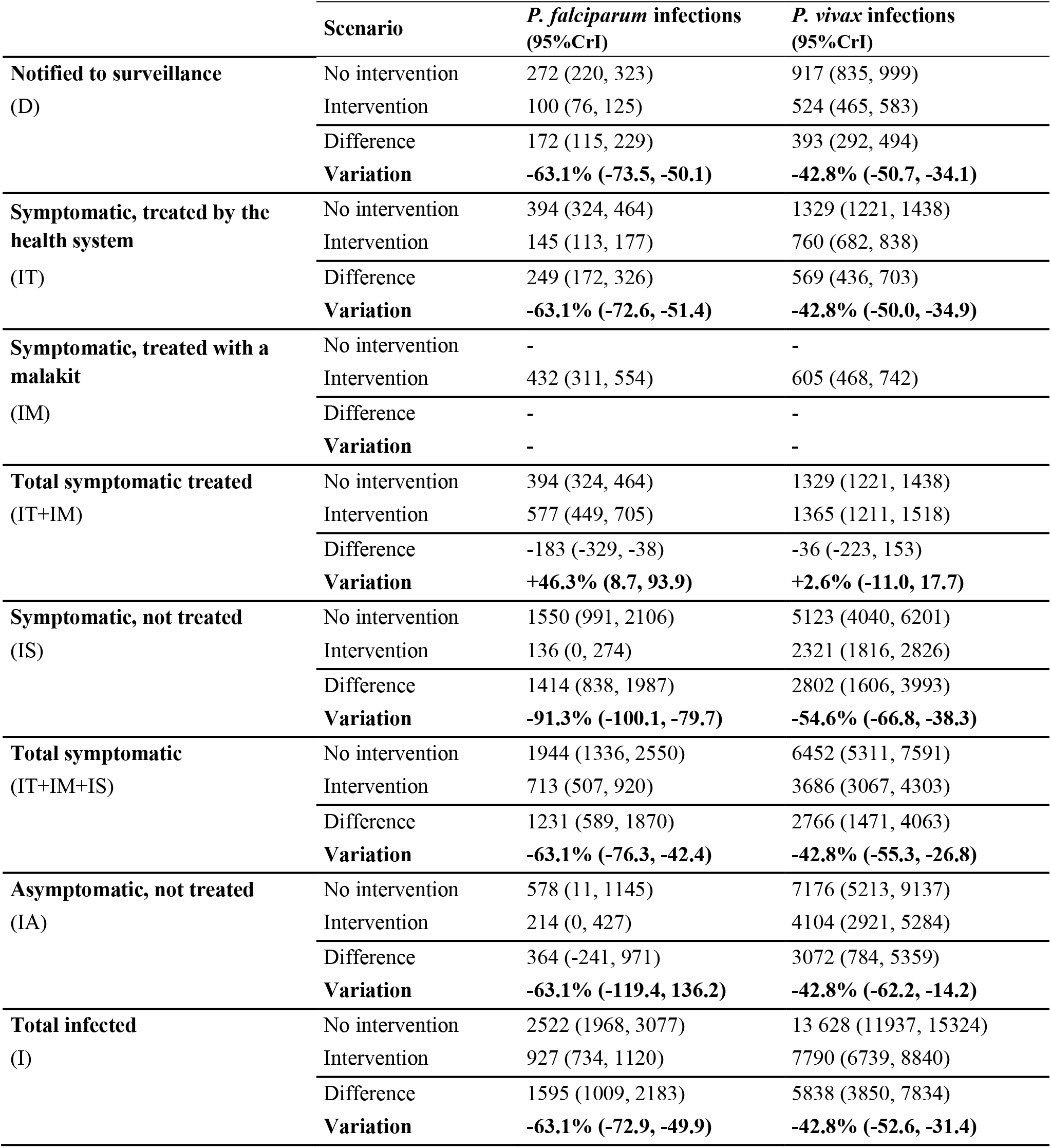
Cumulated malaria incidence of *P. falciparum* and *P. vivax* malaria infections estimated between April 2018 and March 2020, with intervention (model fitted to data) and without (counterfactual)

|  | Scenario | *P. falciparum* infections<br>(95%CrI) | *P. vivax* infections<br>(95%CrI) |
| --- | --- | --- | --- |
| **Notified to surveillance**<br>(D) | No intervention | 272 (220, 323) | 917 (835, 999) |
|  | Intervention | 100 (76, 125) | 524 (465, 583) |
|  | Difference | 172 (115, 229) | 393 (292, 494) |
|  | **Variation** | **-63.1% (-73.5, -50.1)** | **-42.8% (-50.7, -34.1)** |
| **Symptomatic, treated by the health system**<br>(IT) | No intervention | 394 (324, 464) | 1329 (1221, 1438) |
|  | Intervention | 145 (113, 177) | 760 (682, 838) |
|  | Difference | 249 (172, 326) | 569 (436, 703) |
|  | **Variation** | **-63.1% (-72.6, -51.4)** | **-42.8% (-50.0, -34.9)** |
| **Symptomatic, treated with a malakit**<br>(IM) | No intervention | - | - |
|  | Intervention | 432 (311, 554) | 605 (468, 742) |
|  | Difference | - | - |
|  | **Variation** | - | - |
| **Total symptomatic treated**<br>(IT+IM) | No intervention | 394 (324, 464) | 1329 (1221, 1438) |
|  | Intervention | 577 (449, 705) | 1365 (1211, 1518) |
|  | Difference | -183 (-329, -38) | -36 (-223, 153) |
|  | **Variation** | **+46.3% (8.7, 93.9)** | **+2.6% (-11.0, 17.7)** |
| **Symptomatic, not treated**<br>(IS) | No intervention | 1550 (991, 2106) | 5123 (4040, 6201) |
|  | Intervention | 136 (0, 274) | 2321 (1816, 2826) |
|  | Difference | 1414 (838, 1987) | 2802 (1606, 3993) |
|  | **Variation** | **-91.3% (-100.1, -79.7)** | **-54.6% (-66.8, -38.3)** |
| **Total symptomatic**<br>(IT+IM+IS) | No intervention | 1944 (1336, 2550) | 6452 (5311, 7591) |
|  | Intervention | 713 (507, 920) | 3686 (3067, 4303) |
|  | Difference | 1231 (589, 1870) | 2766 (1471, 4063) |
|  | **Variation** | **-63.1% (-76.3, -42.4)** | **-42.8% (-55.3, -26.8)** |
| **Asymptomatic, not treated**<br>(IA) | No intervention | 578 (11, 1145) | 7176 (5213, 9137) |
|  | Intervention | 214 (0, 427) | 4104 (2921, 5284) |
|  | Difference | 364 (-241, 971) | 3072 (784, 5359) |
|  | **Variation** | **-63.1% (-119.4, 136.2)** | **-42.8% (-62.2, -14.2)** |
| **Total infected**<br>(I) | No intervention | 2522 (1968, 3077) | 13 628 (11937, 15324) |
|  | Intervention | 927 (734, 1120) | 7790 (6739, 8840) |
|  | Difference | 1595 (1009, 2183) | 5838 (3850, 7834) |
|  | **Variation** | **-63.1% (-72.9, -49.9)** | **-42.8% (-52.6, -31.4)** |

## 4 Discussion

We have here presented a mathematical model, coupled with Bayesian inference, to jointly analyze three independent datasets: surveillance data from Brazil, from Suriname, and prevalence data from repeated cross-sectional surveys. We find an increase of treatment coverage from 26.4% to 55.1% with the implementation of the Malakit intervention, which resulted in a strong decrease of malaria transmission, and a drop of the estimated reproduction number from 1.19 to 0.86 for all-species malaria. Indeed, access to prompt treatment by ACT and S-PQ (both medication included in Malakit) accelerate the clearance of circulating gametocytes, and thus decrease the duration of the infection for one episode of *Pf* or *Pv* malaria (18). Because Malakit does not offer a radical cure against *Pv* hypnozoites, subsequent *Pv* relapses cannot be prevented. This can explain the lower impact of Malakit estimated on *Pv* transmission, as reported when ACT was implemented in other settings (19). Malakit could also have had an effect on malaria transmission through the increased use of long lasting insecticidal nets (LLINs), by improving access and awareness of this preventive measure among participants (20). However, there is little evidence about the effectiveness of LLINs for malaria prevention in this context given one fifth of gold miners report working at night and the diurnal biting behavior of *An. darlingi* observed in the Guianese rainforest (21,22).

To our knowledge, Malakit was the only intervention implemented at full scale between 2018 and 2020 in the gold mining population, and no significant changes occurred in the management of the health systems of French Guiana, Suriname and Brazil during that period for this population. Our model accounts for seasonal and long-term climate variations with a force of transmission dependent on temperature, and weather records show that temperature and rainfall were quite stable between 2014 and 2020 in French Guiana. Gold mining activity appears to have remained stable over this period. Police operations implemented since 2008 by the French authorities aim to destroy logistical equipment and mining networks, but they are struggling to reduce the number of gold miners. A stable number of police operations were conducted between 2015 and 2019, with the exception of March-April 2017 (23).

We find that, after two years of intervention, Malakit contributed to halving the incidence of malaria infection from 340 to 184 NSI/1000/year. Those estimates are similar to findings from pre-post field surveys: 306 to 142 NSI/1000/year (supplementary material). The field surveys also indicate a similar 2.4-fold increase of treatment coverage (vs. 2.1 with our model), but with a larger post-intervention estimate: 27.3% before intervention and 66.2% after intervention (supplementary Table 3), compared with 26.4% and 55.1%, respectively, found here.

Our results suggest that a significant progression towards malaria elimination was achieved in our study population during the implementation of Malakit, with estimated values of R below unity, the theoretical threshold for malaria elimination (24). We find that the elimination process of *Pf* preceded the intervention (R=0.96) and accelerated during its implementation (R=0.34). This is mirrored by the evolution of the incidence rate of *Pf* infection, compatible with a shift from a low transmission intensity (> 100 cases/1000/year) to a very low transmission intensity (< 100 cases/1000/year) (25). In contrast, for *Pv*, the shift of R below the elimination threshold was concomitant with the intervention.

According to WHO’s framework, the stratification of malaria risk is an important step for planning and achieving malaria elimination (25). This relies on epidemiological and ecological assessments, which is particularly challenging in the case of our population given their high mobility and the difficulties to reach clandestine gold mining areas. We modeled malaria transmission on a single patch with the assumption of homogeneous transmission in all mining sites, our results thus reflect an averaged picture of a more complex and heterogeneous reality. The progressive reduction of malaria incidence will increase this spatial and temporal heterogeneity, making it even more challenging to evaluate future interventions. Along with immunity loss and mobility, this should be taken into consideration in future works, for example in a stochastic or agent-based model (26).

Generating robust evidence about the effectiveness and impact of public health intervention is difficult, especially when working with mobile and hard-to-reach populations. Clustered randomized trials, commonly held as the gold standard design for outcome evaluation, are impractical to implement in this context (27). As an illustration, we only identified two published trials which evaluated a malaria control intervention amongst a hard-to-reach population in the past ten years (28,29). The model proposed here optimally combines different sources of data, each providing partial information, and its findings are in line with those estimated independently from pre-post intervention field surveys. This model-based triangulation approach can be used in similar situations to better inform decision-making towards malaria elimination in especially challenging environments.

## 5 Conclusion

Through a modeling approach, we assess the effectiveness of Malakit as a malaria control intervention in the specific context of the mobile and hard-to-reach population involved in illegal gold mining in French Guiana. Our findings show that Malakit had a significant impact on both *P. falciparum* and *P. vivax* transmission, through an improved access to prompt treatment by ACT and S-PQ for the treatment of symptomatic malaria infections. Building on the efforts of the healthcare community during the past twenty years, Malakit constitutes an additional step towards malaria elimination in the Guiana Shield. This new intervention – training individuals to self-diagnose and self-treat using a free kit – could integrate strategies aiming at eliminating malaria in populations beyond the reach of the health systems.

## Data Availability

All data produced in the present study are available upon reasonable request to the authors

## Contributors

MD, MG, YL, MSM, SV, HH, PM and AS conceived Malakit study and wrote the protocol.

YL, AS, MD, HH and PM accessed and verified the data.

YL, CP, RM developed the model.

YL coded and calibrated the model, analyzed the data and results.

YL, MD, RM and CP wrote the draft of the manuscript.

All authors reviewed the manuscript.

## Data sharing

Monthly aggregated surveillance data, R scripts and MCMC traces are available upon request to the corresponding author (YL).

## Abbreviations

ACT: Artemisinin combined therapy
ITSA: Interrupted time series analysis
LLIN: Long lasting insecticidal net
NSI: New symptomatic infection
NSI/1000/year: New symptomatic infections per 1000 individuals
PCR: Polymerase chain reaction
*Pf*: *Plasmodium falciparum*
*Pv*: *Plasmodium vivax*
RDT: Rapid diagnostic test
S-PQ: Single-dose primaquine

## Acknowledgments

We thank all the facilitators of the Malakit project who contributed to the successful implementation of the intervention, Dr Lise Musset and Dr Yassamine Lazrek from the Institut Pasteur de la Guyane, for PCR prevalence data, and the Ministries of Health of Suriname and Brazil for the sharing of surveillance data.

OML is a staff member of the Pan American Health Organization / World Health Organization (PAHO/WHO). The author alone is responsible for the views expressed in this publication and do not necessarily represent the decisions, policy or views of the PAHO / WHO

