## Supplementary materials for "Modeling the impact of Malakit intervention: one more step towards malaria elimination in the Guiana Shield?"

### 1 Supplementary methods: datasets

#### 1.1 Data from cross-sectional surveys and Malakit inclusions

##### 1.1.1 Prevalence data

Table 1 summarizes the data available from these different field studies. Percentages of infection by *P. falciparum* or *P. vivax* include mixed infections (by both species; 10% of participants in 2015 survey). We chose to use prevalence data from the French-Surinamese border surveys only because they included a larger sample of participants and ensured a consistent pre-analytic handling of blood samples. In addition, the RDT prevalence measured by systematic self-testing of participants during Malakit training in 2018 was converted into PCR prevalence using a sensitivity of 15% and a specificity of 99% (see section 1.1.2).

Table 1: Summary of data available from cross-sectional surveys and Malakit inclusions

|  | **2015** | **2018** | **2019** |
| --- | --- | --- | --- |
| **Data source** | Cross-sectional survey (Suriname) | Malakit inclusions | Cross-sectional survey (Suriname) |
| **Sample size** | 421 | 1188 | 380 |
| **PCR prevalence** | 22.3% | 15.5% * | 5.3% |
| **% *P. falciparum*** | 60% | 26% ** | 15% |
| **% *P. vivax*** | 50% | 76% ** | 85% |
| * Prevalence of positive self-RDT converted into PCR prevalence with Se=15% and Sp=99%  ** Linear interpolation between 2015 and 2019 | | | |

##### 1.1.2 Estimation of 2018 annual prevalence

One step of the training of Malakit participants consisted in performing a self-RDT under the supervision of a facilitator. We collected blood samples on filter paper for a subsample of 62 participants with a positive RDT; 25 of them were positive in PCR which corresponded to a positive predictive value (PPV) of 40.3% (95%CI 28.1, 52.5). The PCR technique used can detect parasitemia with a limit of about one parasite per microliter (1 p/µL) (Snounou *et al.*, 1993).

In 2019, a total 1071 participants attended a training in Malakit distribution sites in Suriname and 21 of them tested positive with a self-RDT, giving an RDT prevalence p_RDT_ of 2.0%. We used this RDT prevalence, the PCR prevalence measured in the 2019 cross-sectional survey in Suriname (p_PCR_=5.3%) and the PPV estimate of the RDT to determine the sensitivity and specificity of the RDT as shown in Table 2. We found a sensitivity of 15.0% (95%CI 12.9, 17.2) and a specificity of 98.8% (95%CI 98.1, 99.4).

Table 2: Calculation of the sensitivity (Se) and specificity (Sp) of Malakit RDT on the subsample of 1071 Malakit participants trained in Suriname in 2019

|  | **Negative**  **RDT** | **Positive**  **RDT** | **Total** |  | **Se** | **15.0%** |
| --- | --- | --- | --- | --- | --- | --- |
| **Negative PCR** | 1002.1 | 12.5 | 1014.6 |  | **Sp** | **98.8%** |
| **Positive PCR** | 47.9 | 8.5 = 21*VPP | 56.4 = 1071*p_PCR_ |  | PPV | 40.3% |
| **Total** | 1050 | 21 | 1071 |  | NPV | 95.4% |

An RDT prevalence can be converted into PCR prevalence using the following equation:

$p_{PCR}= \frac{p_{RDT}-(1-Sp)}{Se-(1-Sp)}$	(1)

In 2018, 3.4% of Malakit participants tested positive with an RDT during trainings (n=40/1188). Using the above equation and the RDT sensibility and specificity calculated above we get a PCR prevalence of 15.5%.

##### 1.1.3 Estimation of the reporting fraction of cases treated by health system

We used pooled data from all cross-sectional surveys to estimate the reporting fraction of cases treated by health system ($\rho$), i.e. the probability for a symptomatic individual infected with malaria in French Guiana and treated by the health system (in either country) to be notified to national surveillance systems of Brazil or Suriname, the sources of data used to fit the model. Among the 1098 participants to all cross-sectional surveys, 605 participants reported that their last episode of malaria had been in French Guiana. Of them, 208 had sought care to a health center: 64 in French Guiana (30.8%) and 144 in Brazil or Suriname (69.2%). We therefore estimate $\rho$=0.69.

##### 1.1.4 External comparison: estimates of incidence rate and treatment behavior

The following data was not used for model calibration and served as an external comparison for the model’s estimates.

In the pre-intervention surveys in Suriname and Brazil, 183 participants reported an episode of malaria symptoms that started in French Guiana within the year before the survey, we thus estimated the mean incidence rate at 306 new symptomatic infections per 1000 individuals per year (NSI/1000/year) (n=183/599; 95%CI 239, 372). Similarly, we estimate the post-intervention incidence rate at 142 NSI/1000/year (n=71/499; 95%CI 61, 224). Table 3 shows the treatment behavior reported for these participants during their episode of malaria in French Guiana.

Table 3: Treatment behavior of participants to cross-sectional surveys with a history of malaria in French Guiana within one year before the survey

|  | **Before intervention** | | **After intervention** | |
| --- | --- | --- | --- | --- |
|  | **n** | **% (95%CI)** | **n** | **% (95%CI)** |
| **Proper treatment** | **50** | **27.3% (15.0, 39.7)** | **47** | **66.2% (52.7, 79.7)** |
| Malakit | - | - | 30 | 42.3% (24.6, 59.9) |
| Health system | 50 | 27.3% (15.0, 39.7) | 17 | 23.9% (3.7, 44.2) |
| **No or improper treatment** | **133** | **72.7% (65.1, 80.3)** | **24** | **33.8% (14.9, 52.7)** |
| **Total** | **183** | **100%** | **71** | **100%** |

#### 1.2 Weather data

Daily records of weather data for 2014-2020 were obtained from four weather stations spanned across the territory of French Guiana (Météo France, 2022). The data comprised daily maximum, minimum and average temperature, daily rainfall and daily maximum, minimum and average relative humidity. We smoothed the variables by calculating a moving average with a window of 30 days. We calculated the median of each weather variables across the four stations because they showed a strong data correlation, suggesting a good spatial homogeneity. All variables were corelated and captured the same seasonality pattern. As an explanatory variable for transmission, we used the daily maximum temperature because it had fewer missing data. Precisely, we used the daily variation of the maximum daily temperature (in °C) around the average calculated between 2014 and 2020 (31.7°C), with seasonal fluctuations between -2.4°C (2.5^th^ percentile) and +2.9°C (97.5^th^ percentile).

### 2 Supplementary methods: model description

#### 2.1 Model compartments and equations

In this study we used a deterministic compartmental model of the transmission of in a single population encompassing all individuals living in illegal gold mining sites in French Guiana. The model is an adaptation of the classic Susceptible-Infectious-Susceptible (SIS) model developed by Ross and Macdonald (Mandal *et al.*, 2011). Given the long period of time of the model simulations, we did not model explicitly the dynamics of the vector population (Anopheles mosquito) and thus the model only included human compartments. The vector dynamics were summarized into a human-to-human transmission rate, as done by other authors (Koella and Antia, 2003; Smith and Ellis McKenzie, 2004; Dudley *et al.*, 2016).

We assume a constant population size (assumption i):

$S+I= N$	(2a)

$I=I_{A}+I_{S}+I_{T}+I_{M}$	(2b)

We assume a homogeneous mixing of the population (assumption ii). The force of infection is temperature-dependent (assumption iii) and is calculated using three key parameters: the transmission rate at average temperature ($\beta_{0}$), the seasonal amplitude ($\alpha$) and the temporal lag of the transmission rate associated to temperature. We assume that the rate of transmission by asymptomatic individuals is lower than that by symptomatic individuals, with a constant ratio $k$ < 1 (assumption v) and that treatment does not affect the transmission rate (assumption x).

The force of transmission is calculated as follows, with $\Delta K$ being the variation of the maximum daily temperature from the 2014-2020 average:

$\Lambda\left( t \right)=\frac{1}{N}\left( kI_{A}+I_{S}+I_{T}+I_{M} \right){\beta_{0}e}^{\alpha\Delta K\left( t-lag \right)}$	(2c)

The progression between compartment S and the other compartments is determined by three parameters, $\varepsilon$ – the probability of a newly infected individual to be symptomatic (assumption iv), $\varphi$ – the probability of a symptomatic individual to be treated (assumption vi) and $m$ – the probability of a symptomatic individual to use a kit for treatment. The duration of infection without treatment $T_{NT}=\frac{1}{\mu_{NT}}$ is the same for asymptomatic and symptomatic individuals (assumption xi) while treatment shortens the duration of infection to $T_{T}=\frac{1}{\mu_{T}}$ (assumption ix).

$\frac{dS}{dt}= -\Lambda S+\mu_{NT}(I_{A}+I_{S})+\mu_{T}(I_{T}+I_{M})$	(2d)

$\frac{dI_{A}}{dt}=\left( 1-\varepsilon\right)\Lambda S-\mu_{NT}I_{A}$	(2e)

$\frac{dI_{S}}{dt}=\varepsilon\left( 1-\varphi\right)\Lambda S-\mu_{NT}I_{S}$	(2f)

$\frac{dI_{T}}{dt}=\varepsilon(\varphi-m)\Lambda S-\mu_{T}I_{T}$	(2g)

$\frac{dI_{M}}{dt}=\varepsilon m \Lambda S-\mu_{T}I_{M}$	(2h)

Assuming that individuals who use a kit for treatment are not notified (assumption xiii), the incidence rate of notification $r_{D}$ is derived from the the incidence rate of infections treated by the health system$r_{T}$, with $\rho$ being the probability for a symptomatic individual infected in French Guiana and treated by the health system (in either country) to be notified to national surveillance systems of Brazil or Suriname (assumption xii):

$r_{D}=\rho r_{T}$	(2i)

$r_{D}=\rho\varepsilon(\varphi-m)\Lambda S$	(2j)

We define $\varphi_{0}$ and $\varphi_{1}$ as the probabilities for a symptomatic individual to get a treatment prior and during the intervention, respectively. Similarly, $m_{0}$and $m_{1}$ are the probabilities for a symptomatic individual to use a kit for malaria treatment prior and during the intervention, respectively. By definition:

$m_{0}= 0$	(2k)

We assume that the proportion of individuals who seek care from the health remains the same during the intervention period (assumption viii), i.e. the change on care seeking and treatment behavior associated to the intervention occurred only in the population who did not have access to the health system:

$\varphi_{0}=\varphi_{1}-m_{1}$	(2l)

We can then calculate the probability of using a kit as a treatment during intervention as follows:

$m_{1}= \varphi_{1}-\varphi_{0}$	(2m)

We define the reproduction number at average temperature as follows:

$R=\left( 1-\varepsilon\right)\frac{{k\beta}_{0}}{\mu_{NT}}+ \varepsilon\left( 1-\varphi\right)\frac{\beta_{0}}{\mu_{NT}}+\varepsilon\varphi\frac{\beta_{0}}{\mu_{T}}$	(2n)

We define the mean infection duration (for all infections, irrespective of treatment) as follows:

$T=\frac{R}{\beta_{0}}$	(2o)

#### 2.2 Parameter definitions et estimation methods

Table 4: Parameters notation, definition and method of estimation. u(x,y): uniform distribution. Se: values used in sensitivity analysis

| Parameter description | Notation | Values/prior distribution | Source/Estimated |
| --- | --- | --- | --- |
| *Natural history of disease and demographics* | | | |
| Total population size | $N$ | 10 000 | (Douine *et al.*, 2017, p. 201; Heemskerk, Eelco Jacobs, and Pierre Pratley, 2021) |
| Number of infected individuals at t_0_,  for all species | $I_{0}$ | 3000 (Se: 4000) | (Douine *et al.*, 2016; Pommier de Santi *et al.*, 2016) |
| Number of infected individuals at t_0_,  for *P. falciparum* | $I_{0,pf}$ | 2100 (Se: 2800) |  |
| Number of infected individuals at t_0_,  for *P. vivax* | $I_{0,pv}$ | 1650 (Se: 2200) |  |
| Proportion of symptomatic individuals at t_0_ | $\frac{I_{S,0}}{I_{0}}$ | 0.50 | (Pommier de Santi *et al.*, 2016) |
| Infection duration without treatment (day), for *P. falciparum* | $T_{NT,pf}=\frac{1}{\mu_{NT,pf}}$ | 160 (Se: 140, 180) | (Sama *et al.*, 2005; Felger *et al.*, 2012; Bretscher *et al.*, 2015) |
| Infection duration without treatment (day), for *P. vivax* | $T_{NT, pv}=\frac{1}{\mu_{NT, pv}}$ | 70 (Se: 60, 80) | (Kerlin and Gatton, 2015) |
| Infection duration without treatment (day), for all species | $T_{NT}=\frac{1}{\mu_{NT}}$ | 100 (Se: 85, 110) | $\mu_{NT}=mean(\mu_{NT, pf}, \mu_{NT,pv})$ |
| Infection duration with treatment (day),  for all species | $T_{T}=\frac{1}{\mu_{T}}$ | 7 | (Bousema *et al.*, 2010; White, 2013; Eziefula *et al.*, 2014) |
| Mean infection duration (all infections, irrespective of treatment) | $T$ | - | Equation 2o |
| *Transmission-related parameters* | | | |
| Force of infection | $\Lambda$ | - | Equation 2c |
| Transmission rate at average maximum daily temperature (day^-1^) | $\beta_{0}$ | u(0.001, 0.050) | Estimated by fitting model to data |
| Temperature-associated amplitude of transmission rate (°C^-1^) | $\alpha$ | u(0, 0.5) | Estimated by fitting model to data |
| Temperature-associated lag of transmission rate (day) | $lag$ | u(0, 360) | Estimated by fitting model to data |
| Variation from 2014-2020 average maximum daily temperature (°C) | $\Delta K$ | - | (Météo France, 2022), see section 1.2 |
| Symptomatic/asymptomatic transmission rate ratio, for *P. falciparum* | $k_{pf}$ | 0.33 | (Slater *et al.*, 2019) |
| Symptomatic/asymptomatic transmission rate ratio, for *P. vivax* | $k_{pv}$ | 0.10 | (Ferreira *et al.*, 2022) |
| Symptomatic/asymptomatic transmission rate ratio, for all species | $k$ | 0.18 | Geometric mean of $k_{pf}$ and $k_{pv}$ |
| Reproduction number at average temperature | $R$ | - | Equation 2n |
| *Symptoms, treatment and case reporting parameters* | | | |
| Probability of symptomatic infection | $\varepsilon$ | u(0, 1) | Estimated by fitting model to data |
| Probability for a symptomatic individual to get treatment, prior intervention | $\varphi_{0}$ | u(0, 1) | Estimated by fitting model to data |
| Probability for a symptomatic individual to get treatment, during intervention | $\varphi_{1}$ | u(0, 1) | Estimated by fitting model to data |
| Probability for a symptomatic individual to use a kit, prior intervention | $m_{0}$ | 0 | Equation 2k |
| Probability for a symptomatic individual to use a kit, during intervention | $m_{1}$ | - | Equation 2m |
| Reporting fraction of cases treated by health system | $\rho$ | 0.69 | See section 1.1.3 |

#### 2.3 Model fitting and parameter estimation

##### 2.3.1 Log-likelihood calculation

We fitted the simulated monthly malaria notification incidence $r_{D,m}$ to the monthly number of notified cases of malaria $x_{m}$, and we fitted the simulated mean annual prevalence of infectious cases $p_{I,y}$ to the number of infected individuals $i_{y}$ measured in 2015, 2018 and 2019 prevalence surveys. The number of cases notified in a given month $m$ followed a Poisson distribution and the number of infected individuals measured in a prevalence survey of a given year $y$on a sample of $n_{y}$ individuals followed a binomial distribution. The corresponding log-likelihoods ${LogLik}_{m}$ and ${LogLik}_{y}$ and the overall log-likelihood ${LogLik}_{all}$are presented in the following equations.

$x_{m}\sim Pois\left( {rD}_{m} \right)$	(3a)

$i_{y}\sim Bin\left( n_{y}, {pI}_{y} \right)$	(3b)

${LogLik}_{m}\left( x_{m} | \theta\right)=x_{m}\log\left( r_{D,m}\left( \theta\right) \right)-r_{D,m}\left( \theta\right)-log(x_{m}!)$	(3c)

${LogLik}_{y}\left( i_{y},n_{y} | \theta\right)=i_{y}\log\left( p_{I,y}\left( \theta\right) \right)+\left( n_{y}-i_{y} \right)\log\left( 1-p_{I,y}\left( \theta\right) \right)$	(3d)

${LogLik}_{all}\left( data | \theta\right)=\sum_{m} {LogLik}_{m}\left( x_{m} | \theta\right)+\sum_{y} {LogLik}_{y}\left( i_{y} | \theta\right)$	(3e)

##### 2.3.2 Credible interval calculation

We ran 1000 simulations (noted *i*) of the model, using a random sample of 1000 draws from the joint posterior distributions of the parameters. This resulted in 1000 model trajectories, giving for each month *m* (time step) a distribution of 1000 $r_{m,i}$values for the estimated monthly incidence rate $r_{m}$.

We assumed that this distribution was normal with mean $\bar{r_{m}}=mean\left( r_{m,i} \right)$ and variance $\sigma^{2}$ :

$r_{m}\mathcal{\sim N}\left( \bar{r_{m}},\sigma^{2} \right)$	(4a)

In addition, to account for the uncertainty resulting from the observational process in the mean value $\bar{r_{m}}$of the simulated incidence rate at each month, we assumed that each one of the 1000 simulated $r_{m,i}$values followed a Poisson distribution of rate $r_{m,i}$.

$r_{m,i} \sim Pois(r_{m,i})$	(4b)

We approximated the sum of these 1000 Poisson distributions with a normal distribution of mean $\bar{r_{m}}$ and variance $\bar{r_{m}}$:

$\sum_{i} Pois(r_{m,i})\mathcal{\approx N}\left( \bar{r_{m}}, \bar{r_{m}} \right)$	(4c)

We then determined the full distribution of the monthly incidence rate $r_{m}$ by combining the two distributions (4a) and (4c) described above, resulting in the following approximate normal distribution:

$r_{m}\mathcal{\sim N}\left( \bar{r_{m}},\sigma^{2} \right)+ \sum_{i} Pois(r_{m,i})\mathcal{\approx N}\left( \bar{r_{m}},\sigma^{2}+\bar{r_{m}} \right)$	(4d)

When computing the impact of Malakit, we used the *rnorm* function of R to simulate the above distribution of cumulated incidence and annual incidence rates for the data-fit and counterfactual scenarios, as well as the distribution of the difference and the distribution of the % variation between the two scenarios. We then computed the corresponding 95%CrIs from these distributions.

##### 2.3.3 Configuration of MCMC

For each model fit we ran three Markov chains of 100 000 iterations using different seed values, as shown in Table 5.

Table 5: Seed values used for the estimated parameters, for each of the three Markov chains

| **Parameter** | **Chain #1** | **Chain #2** | **Chain #3** |
| --- | --- | --- | --- |
| Alpha | 0.1 | 0.3 | 0.2 |
| Lag | 100 | 60 | 80 |
| Epsilon | 0.3 | 0.5 | 0.7 |
| Beta0 | 0.020 | 0.015 | 0.010 |
| Phi0 | 0.3 | 0.2 | 0.1 |
| Phi1 | 0.8 | 0.5 | 0.4 |

### 3 Supplementary results

#### 3.1 Main results

Table 6: Estimated impact of the Malakit intervention on mean incidence rate (new symptomatic infections per 1000 individuals per year) and total prevalence (%, all infections) for *P. falciparum* (Pf), *P. vivax* (Pv) and both species (mean and 95% credible interval).

|  |  | **Incidence rate** | | | | **Total prevalence** | | |
| --- | --- | --- | --- | --- | --- | --- | --- | --- |
|  |  | ***Pf*** | ***Pv*** | **All** | ***Pf*** | | ***Pv*** | **All** |
| **B** | Baseline (pre-intervention) | 153 (116, 191) | 340 (283, 397) | 366 (304, 428) | 9.0 (8.0, 10.2) | | 12.7 (11.1, 14.3) | 19.6 (17.8, 21.4) |
| **C** | Counterfactual (no intervention) | 97 (62, 133) | 323 (256, 389) | 330 (262, 398) | 5.0 (4.0, 6.2) | | 12.0 (10.4, 13.6) | 17.1 (15.2, 19.2) |
| **I** | Intervention | 36 (20, 51) | 184 (144, 225) | 169 (131, 207) | 2.2 (1.7, 2.8) | | 7.2 (6.2, 8.2) | 9.6 (8.5, 10.7) |
| **C vs. B** | Difference from baseline (without intervention) | 56 (4, 108) | 17 (-70, 105) | 36 (-56, 128) | 4.0 (3.3, 4.7) | | 0.7 (0.1, 1.3) | 2.5 (1.7, 3.2) |
|  | Variation from baseline (without intervention) | -36.9% (-61.6, -3.4) | -5.2% (-28.1, 23.4) | -10.0% (-31.7, 17.3) | -44.1% (-51.8, -36.3) | | -5.8% (-10.2, -1.1) | -12.6% (-16.8, -8.7) |
| **I vs. B** | Difference from baseline (with intervention) | 118 (77, 158) | 156 (86, 226) | 197 (124, 270) | 6.8 (6.2, 7.5) | | 5.5 (4.7, 6.4) | 10.0 (9.0, 11.0) |
|  | Variation from baseline (with intervention) | -76.8% (-87.2, -63.2) | -45.8% (-59.5, -28.9) | -53.8% (-65.7, -39.1) | -75.6% (-79.0, -72.0) | | -43.1% (-46.8, -39.1) | -51.0% (-53.5, -48.2) |
| **I vs. C** | Difference | 62 (23, 100) | 138 (61, 216) | 161 (83, 238) | 2.8 (2.1, 3.7) | | 4.7 (3.9, 5.7) | 7.5 (6.4, 8.8) |
|  | Variation | -63.1% (-80.6, -34.6) | -42.8% (-58.1, -22.7) | -48.7% (-62.6, -30.4) | -56.2% (-62.9, -49.1) | | -39.6% (-44.0, -34.9) | -43.9% (-47.5, -40.2) |
| **(C-I)/**  **(B-I)** | **Contribution of the intervention to total variation from baseline** | 52.0% | 88.6% | 81.4% | 41.7% | | 86.7% | 75.4% |

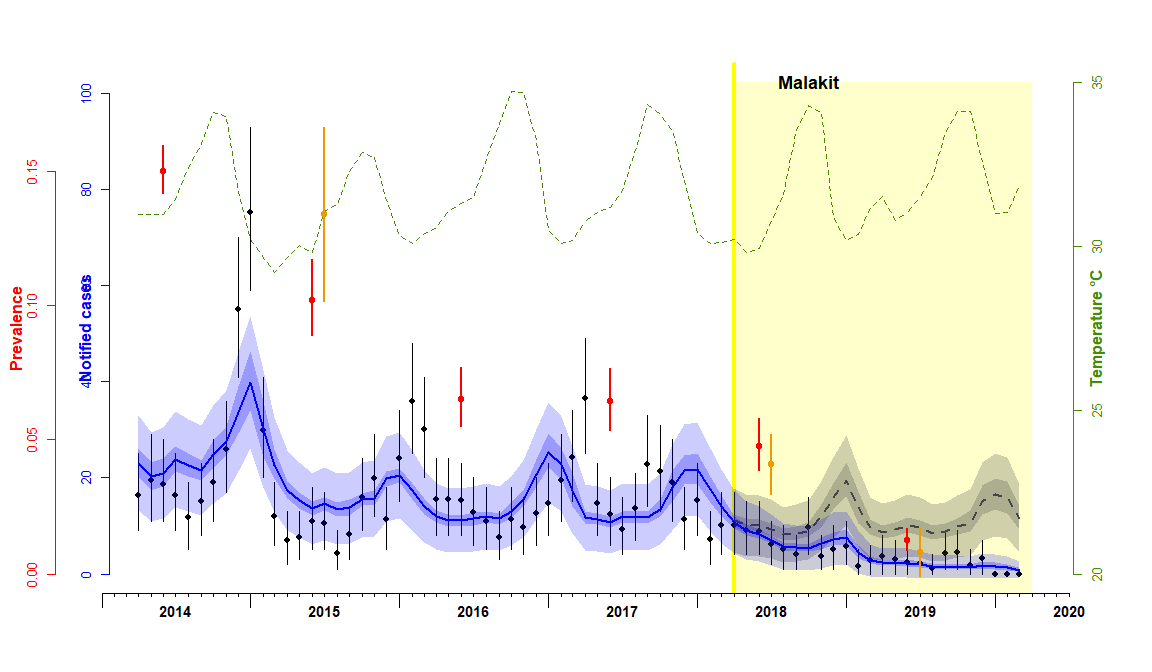

Figure 1: Monthly incidence of *P. falciparum* reported cases of malaria (black dots), model-fitted simulations (blue solid line) and counterfactual estimates (dashed grey line). Annual all-species prevalence measured in PCR surveys (orange dots) and model-fitted estimates of prevalence (red dots). Maximum daily temperature (monthly average) is shown in green dashed line. The period of implementation of Malakit is shown in yellow. Vertical lines represent 95%CrI. For model-fitted and counterfactual incidence (respectively blue and grey), the dark envelopes show the uncertainty (95%CrI) associated to the joint posterior distribution, while the light envelopes account also for the uncertainty resulting from of the observed Poisson process. (T_NT_=160 days, I_0_=2100)

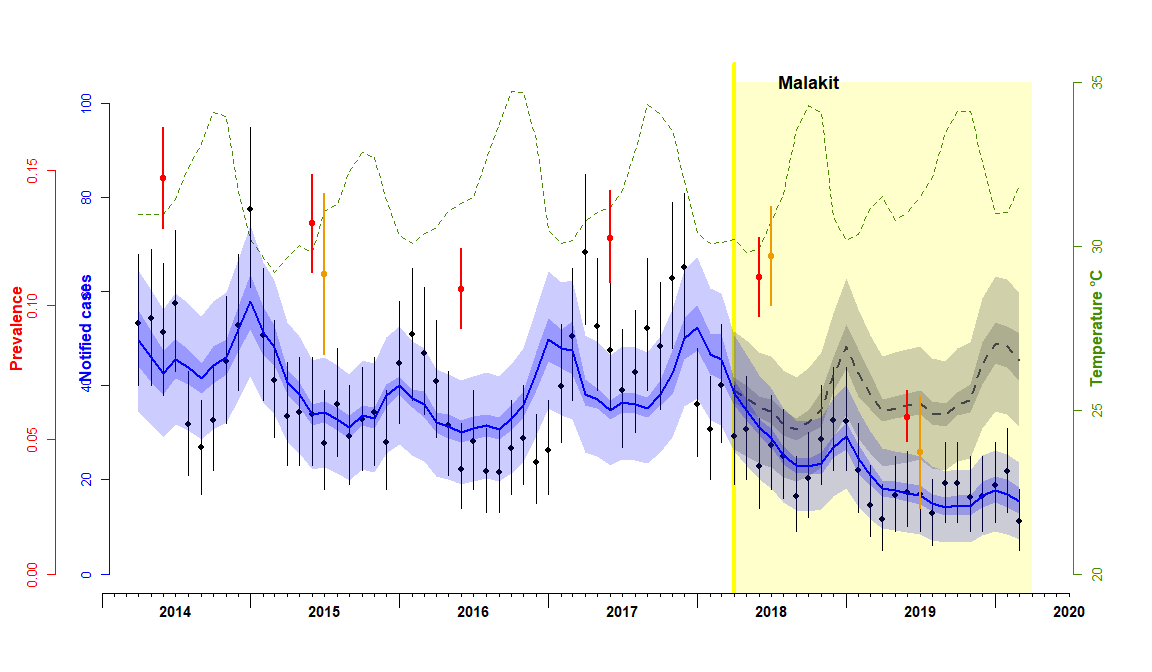

Figure 2: Monthly incidence of *P. vivax* reported cases of malaria (black dots), model-fitted simulations (blue solid line) and counterfactual estimates (dashed grey line). Annual all-species prevalence measured in PCR surveys (orange dots) and model-fitted estimates of prevalence (red dots). Maximum daily temperature (monthly average) is shown in green dashed line. The period of implementation of Malakit is shown in yellow. Vertical lines represent 95%CrI. For model-fitted and counterfactual incidence (respectively blue and grey), the dark envelopes show the uncertainty (95%CrI) associated to the joint posterior distribution, while the light envelopes account also for the uncertainty resulting from of the observed Poisson process. (T_NT_=70 days, I_0_=1650)

#### 3.2 Sensitivity analysis

We assessed the sensitivity of parameter estimation to the value of T_NT_ (duration of infection without treatment) and I_0_ (number of individuals infected at t=0). As shown in Table 4, we fitted the model with three different fixed values of T_NT_ and two different fixed values of I_0_:

- T_NT_ = 85, 100 and 110 days; I_0_ = 3000 and 4000 for all-species model
- T_NT_ = 140, 160 and 180 days; I_0_ = 2100 and 2800 for *P. falciparum* model
- T_NT_ = 60, 70 and 80 days; I_0_ = 1650 and 2200 for *P. vivax* model

The posterior distributions of the parameters estimated for the different combinations of T_NT_ and I_0_ values are shown in Table 7 (all-species), Table 8 (*P. falciparum*) and Table 9 (*P. vivax*). The effect of a variation of T_NT_ and I_0_ was modest on the estimation of m (probability of using a malakit as treatment when symptomatic), α (temperature-associated amplitude of the transmission rate) and the lag of the transmission rate. The estimates of R_0_ and R_1_, the reproduction number prior and during intervention, showed little or no variation with either T_NT_ or I_0_ mainly because their variation was compensated by that of the transmission rate β_0_ which decreased with higher values of T_NT_ and I_0_.

The estimates of φ_0_ and φ_1_ increased with the value of T_NT_ and decreased with I_0_, but with consistently overlapping 95% credible intervals: in the all-species model, φ_0_ and φ_1_ varied from 0.22 (95%CrI 0.19, 0.25) and 0.50 (95%CrI 0.46, 0.55) with T_NT_=100 days and I_0_=4000, to respectively 0.28 (95%CrI 0.24, 0.32) and 0.57 (95%CrI 0.52, 0.63), with T_NT_=110 days and I_0_=3000.

The parameter most sensitive to a variation of I_0_ was ε (the probability of symptomatic infection), however with consistently overlapping 95% credible intervals: for example, the estimate of ε for the all-species model varied between 0.47 (95%CrI 0.39, 0.57) and 0.57 (95%CrI 0.47, 0.67) for I_0_ set to 3000 and 4000, respectively, and T_NT_=100 days.

Table 7: Posterior distributions of model parameters, all-species (mean and 95%CrI)

| **Parameter** | **I_0_** | **T_NT_ = 85** | **T_NT_ = 100** | **T_NT_ = 110** |
| --- | --- | --- | --- | --- |
| **φ_0_** | 3000 | 0.24 (0.21, 0.28) | 0.26 (0.23, 0.30) | 0.28 (0.24, 0.32) |
|  | 4000 | - | 0.22 (0.19, 0.25) | - |
| **φ_1_** | 3000 | 0.52 (0.47, 0.58) | 0.55 (0.50, 0.61) | 0.57 (0.52, 0.63) |
|  | 4000 | - | 0.50 (0.46, 0.55) | - |
| **m** | 3000 | 0.28 (0.24, 0.32) | 0.29 (0.25, 0.33) | 0.30 (0.26, 0.33) |
|  | 4000 | - | 0.28 (0.25, 0.32) | - |
| **ε** | 3000 | 0.45 (0.36, 0.54) | 0.47 (0.39, 0.57) | 0.49 (0.40, 0.57) |
|  | 4000 | - | 0.57 (0.47, 0.67) | - |
| **β_0_** | 3000 | 0.0316 (0.0270, 0.0371) | 0.0265 (0.0226, 0.0308) | 0.0240 (0.0207, 0.0277) |
|  | 4000 | - | 0.0226 (0.0194, 0.0261) | - |
| **α** | 3000 | 0.092 (0.075, 0.109) | 0.097 (0.078, 0.115) | 0.100 (0.081, 0.119) |
|  | 4000 | - | 0.097 (0.078, 0.116) | - |
| **lag** | 3000 | 81 (71, 91) | 83 (73, 94) | 84 (75, 95) |
|  | 4000 | - | 84 (74, 94) | - |
| **T_0_** | 3000 | 38 (33, 44) | 45 (39, 52) | 50 (43, 57) |
|  | 4000 | - | 53 (46, 61) | - |
| **T_1_** | 3000 | 29 (24, 33) | 33 (28, 38) | 35 (30, 40) |
|  | 4000 | - | 38 (33, 44) | - |
| **R_0_** | 3000 | 1.20 (1.17, 1.23) | 1.19 (1.16, 1.22) | 1.19 (1.16, 1.22) |
|  | 4000 | - | 1.20 (1.17, 1.23) | - |
| **R_1_** | 3000 | 0.90 (0.87, 0.93) | 0.86 (0.83, 0.90) | 0.84 (0.80, 0.87) |
|  | 4000 | - | 0.86 (0.83, 0.89) | - |

Table 8: Posterior distributions of model parameters, *P. falciparum* (mean and 95%CrI)

| **Parameter** | **I_0_** | **T_NT_ = 140** | **T_NT_ = 160** | **T_NT_ = 180** |
| --- | --- | --- | --- | --- |
| **φ_0_** | 2100 | 0.19 (0.15, 0.24) | 0.21 (0.17, 0.26) | 0.22 (0.19, 0.27) |
|  | 2800 | - | 0.17 (0.14, 0.20) | - |
| **φ_1_** | 2100 | 0.75 (0.62, 0.91) | 0.82 (0.68, 0.96) | 0.87 (0.73, 0.99) |
|  | 2800 | - | 0.75 (0.63, 0.89) | - |
| **m** | 2100 | 0.56 (0.44, 0.70) | 0.61 (0.48, 0.74) | 0.65 (0.51, 0.76) |
|  | 2800 | - | 0.59 (0.46, 0.72) | - |
| **ε** | 2100 | 0.75 (0.53, 0.96) | 0.77 (0.56, 0.97) | 0.80 (0.60, 0.98) |
|  | 2800 | - | 0.90 (0.72, 1.00) | - |
| **β_0_** | 2100 | 0.0101 (0.0085, 0.0122) | 0.0087 (0.0074, 0.0104) | 0.0076 (0.0066, 0.0090) |
|  | 2800 | - | 0.0076 (0.0070, 0.0087) | - |
| **α** | 2100 | 0.17 (0.13, 0.20) | 0.17 (0.14, 0.21) | 0.18 (0.14, 0.22) |
|  | 2800 | - | 0.18 (0.14, 0.21) | - |
| **lag** | 2100 | 76 (64, 87) | 78 (66, 89) | 79 (67, 90) |
|  | 2800 | - | 78 (67, 89) | - |
| **T_0_** | 2100 | 98 (80, 116) | 112 (91, 131) | 126 (105, 145) |
|  | 2800 | - | 127 (109, 137) | - |
| **T_1_** | 2100 | 42 (30, 56) | 40 (26, 56) | 36 (20, 54) |
|  | 2800 | - | 46 (29, 63) | - |
| **R_0_** | 2100 | 0.98 (0.95, 1.01) | 0.96 (0.93, 1.00) | 0.95 (0.91, 0.98) |
|  | 2800 | - | 0.96 (0.93, 0.99) | - |
| **R_1_** | 2100 | 0.42 (0.31, 0.53) | 0.34 (0.22, 0.46) | 0.27 (0.15, 0.40) |
|  | 2800 | - | 0.35 (0.22, 0.47) | - |

Table 9: Posterior distributions of model parameters, *P. vivax* (mean and 95%CrI)

| **Parameter** | **I_0_** | **T_NT_ = 60** | **T_NT_ = 70** | **T_NT_ = 80** |
| --- | --- | --- | --- | --- |
| **φ_0_** | 1650 | 0.19 (0.16, 0.21) | 0.21 (0.18, 0.24) | 0.22 (0.20, 0.25) |
|  | 2200 | - | 0.17 (0.14, 0.19) | - |
| **φ_1_** | 1650 | 0.34 (0.31, 0.37) | 0.37 (0.34, 0.41) | 0.40 (0.36, 0.43) |
|  | 2200 | - | 0.33 (0.30, 0.37) | - |
| **m** | 1650 | 0.16 (0.13, 0.18) | 0.16 (0.14, 0.19) | 0.17 (0.14, 0.20) |
|  | 2200 | - | 0.17 (0.14, 0.19) | - |
| **ε** | 1650 | 0.46 (0.39, 0.56) | 0.47 (0.38, 0.59) | 0.50 (0.41, 0.60) |
|  | 2200 | - | 0.59 (0.44, 0.71) | - |
| **β_0_** | 1650 | 0.0431 (0.0356, 0.0494) | 0.0371 (0.0300, 0.0451) | 0.0317 (0.0263, 0.0377) |
|  | 2200 | - | 0.0300 (0.0247, 0.0389) | - |
| **α** | 1650 | 0.069 (0.052, 0.086) | 0.075 (0.057, 0.093) | 0.079 (0.060, 0.098) |
|  | 2200 | - | 0.075 (0.057, 0.093) | - |
| **lag** | 1650 | 80 (66, 94) | 82 (68, 97) | 84 (70, 99) |
|  | 2200 | - | 82 (68, 97) | - |
| **T_0_** | 1650 | 26 (23, 31) | 31 (25, 37) | 36 (30, 43) |
|  | 2200 | - | 38 (30, 45) | - |
| **T_1_** | 1650 | 23 (20, 27) | 26 (21, 31) | 30 (25, 35) |
|  | 2200 | - | 32 (25, 38) | - |
| **R_0_** | 1650 | 1.13 (1.11, 1.15) | 1.13 (1.11, 1.15) | 1.13 (1.11, 1.15) |
|  | 2200 | - | 1.13 (1.11, 1.15) | - |
| **R_1_** | 1650 | 0.97 (0.95, 0.99) | 0.95 (0.92, 0.98) | 0.93 (0.90, 0.96) |
|  | 2200 | - | 0.95 (0.92, 0.97) | - |

#### 3.3 MCMC-MH

##### 3.3.1 Diagnoses

As shown in Table 9, the acceptance rates (AR) of the Metropolis-Hastings algorithm are consistently between 22% and 25%, i.e. close to the value of 23.4% which is commonly considered as optimal. Conversely, the Gelman multivariate potential scale reduction factors are close to 1.000, suggesting a good convergence of the three Markov chains run for each model as shown graphically in the corresponding trace plots in sections 3.3.2 to 3.3.4. The deviance information criteria (DIC) do not vary significantly between the different variations of the sensitivity analysis and, within a given model, between the three Markov chains.

The plots shown in Figure 5, Figure 8 and Figure 11 indicate a strong correlation between four estimated parameters (ε, β_0_, φ_0_, φ_1_) which did not impact the mixing and convergence of the MCMC-MH algorithm.

Table 10: MCMC-MH diagnoses for the three main models (in bold) and their respective variations used in the sensitivity analysis. AR: acceptance rate, DIC: deviance information criterion, mPSRF: multivariate potential scale reduction factor.

| **Species** | **T_NT_** | **I_0_** | **Chain #1** | | **Chain #2** | | **Chain #3** | | **Gelman mPSRF** |
| --- | --- | --- | --- | --- | --- | --- | --- | --- | --- |
|  |  |  | **AR** | **DIC** | **AR** | **DIC** | **AR** | **DIC** |  |
| **All** | **100** | **3000** | **0.232** | **697.0** | **0.242** | **697.5** | **0.240** | **697.4** | **1.0023** |
|  | 85 | 3000 | 0.237 | 696.3 | 0.244 | 695.2 | 0.239 | 697.3 | 1.0057 |
|  | 110 | 3000 | 0.239 | 697.5 | 0.238 | 698.1 | 0.238 | 696.6 | 1.0022 |
|  | 100 | 4000 | 0.236 | 695.6 | 0.237 | 697.2 | 0.234 | 695.3 | 1.0040 |
| **Pf** | **160** | **2100** | **0.240** | **550.6** | **0.233** | **549.3** | **0.238** | **550.9** | **1.0022** |
|  | 140 | 2100 | 0.244 | 553.8 | 0.231 | 554.3 | 0.231 | 553.0 | 1.0024 |
|  | 180 | 2100 | 0.245 | 547.6 | 0.240 | 546.6 | 0.232 | 546.3 | 1.0023 |
|  | 160 | 2800 | 0.254 | 549.0 | 0.230 | 549.8 | 0.241 | 549.2 | 1.0030 |
| **Pv** | **70** | **1650** | **0.253** | **589.3** | **0.258** | **584.3** | **0.228** | **591.5** | **1.0163** |
|  | 60 | 1650 | 0.232 | 582.6 | 0.251 | 579.6 | 0.235 | 582.1 | 1.0043 |
|  | 80 | 1650 | 0.250 | 586.8 | 0.230 | 585.5 | 0.238 | 590.1 | 1.0274 |
|  | 70 | 2200 | 0.229 | 600.7 | 0.227 | 591.8 | 0.229 | 585.6 | 1.0038 |

##### 3.3.2 All-species model

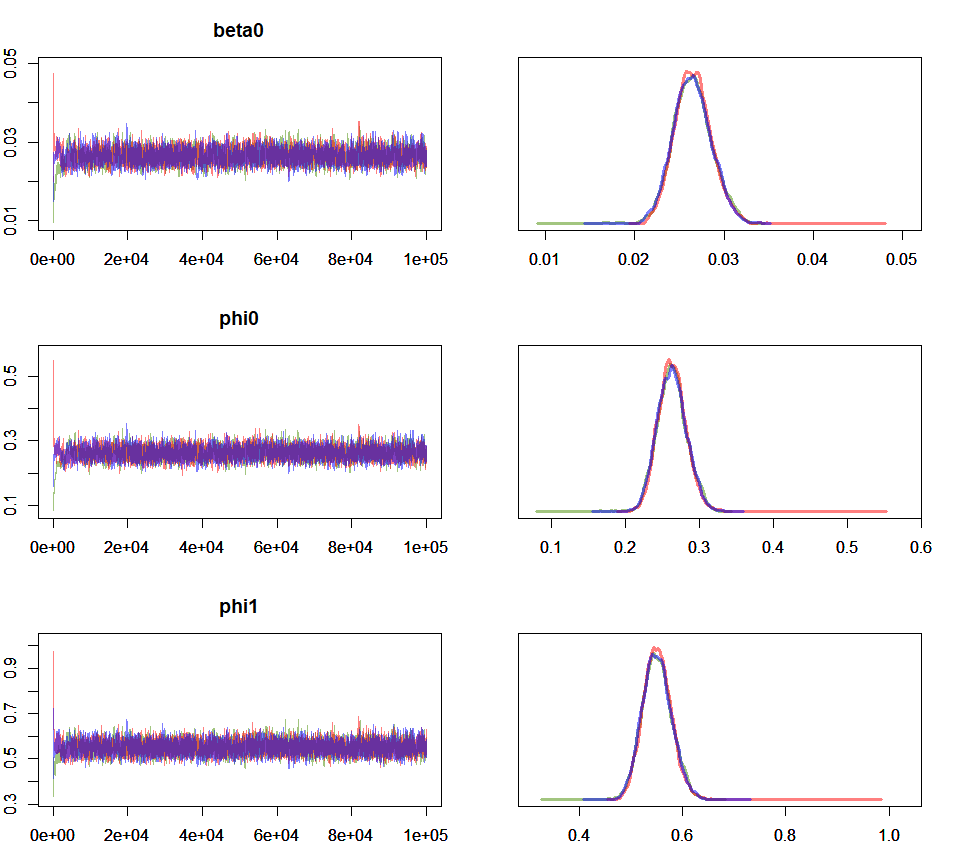

Figure 3: Trace and density plots for parameters beta0, phi0 and phi1. Model fitted to all species data with T_NT_=100 days and I_0_=3000. Red: chain #1, Blue: chain #2, Green: chain #3.

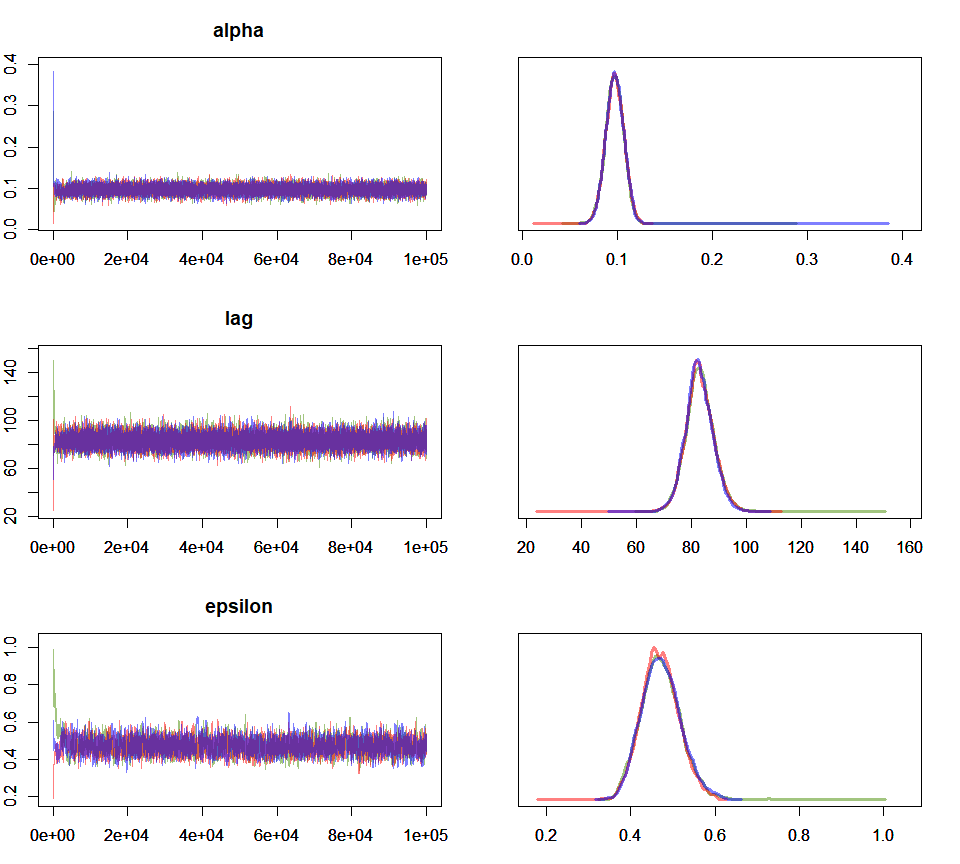

Figure 4: Trace and density plots for parameters alpha, lag and epsilon. Model fitted to all species data with T_NT_=100 days and I_0_=3000. Red: chain #1, Blue: chain #2, Green: chain #3.

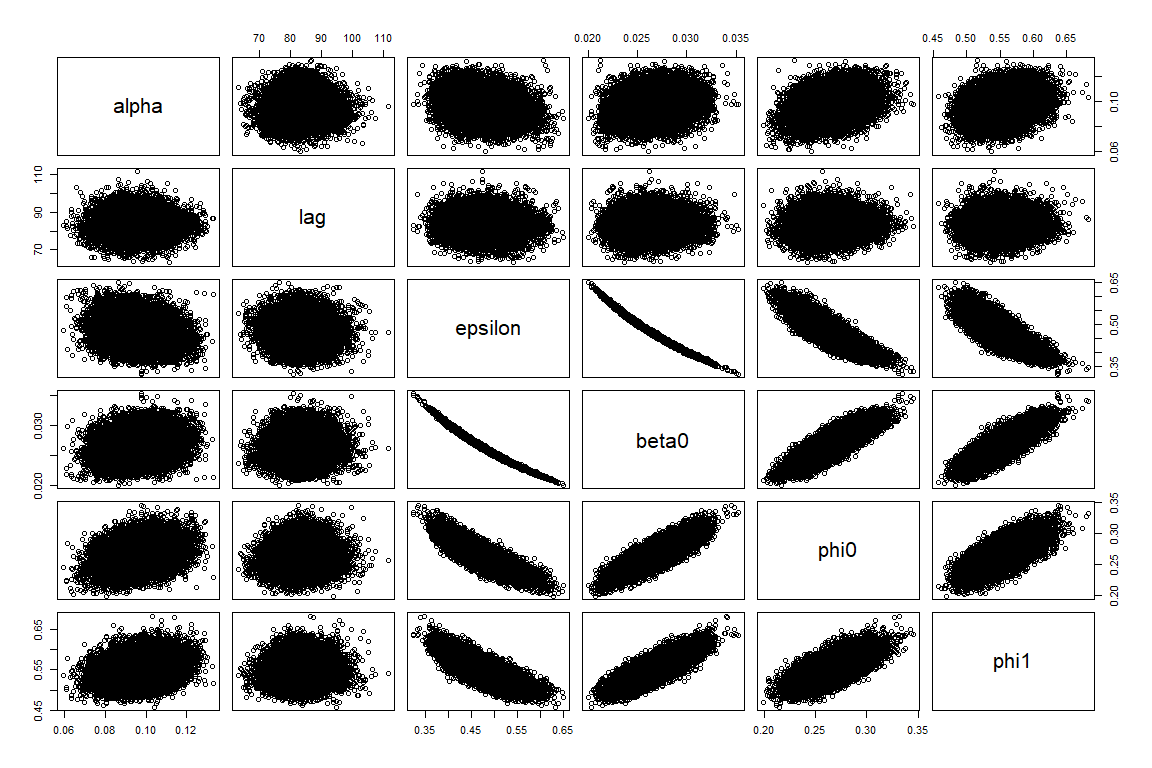

Figure 5: Correlation plots, model fitted to all species data with T_NT_=100 days and I_0_=3000.

##### 3.3.3 *P. falciparum* model

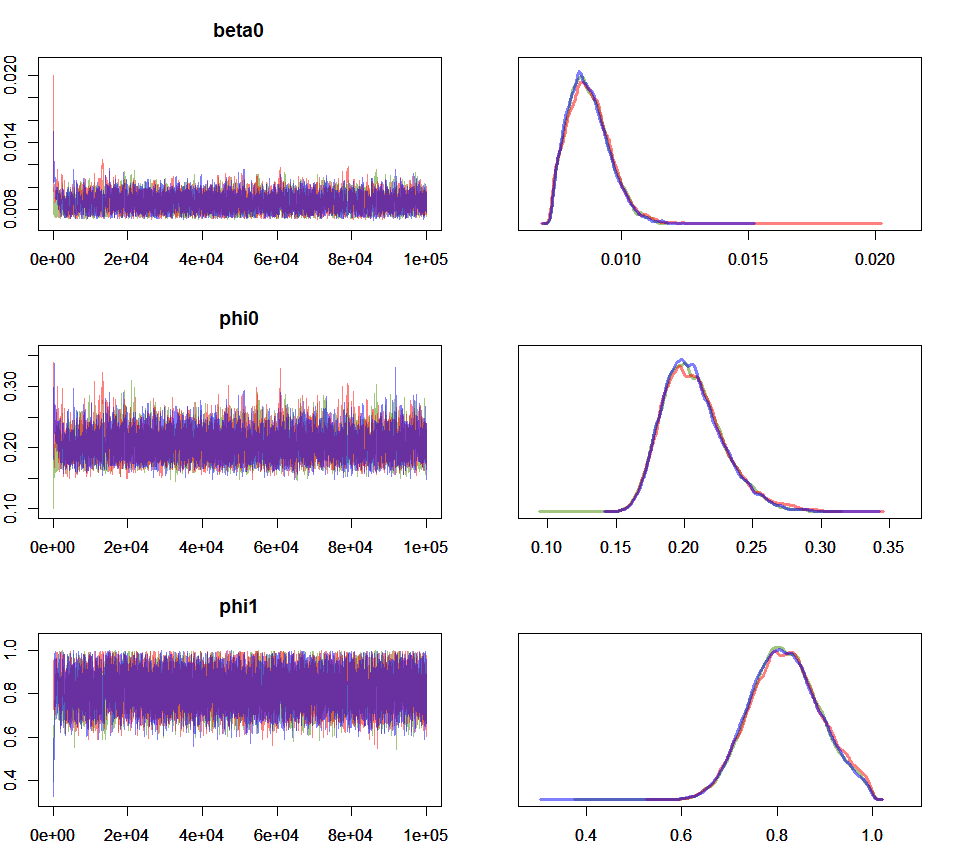

Figure 6: Trace and density plots for parameters beta0, phi0 and phi1. Model fitted to *P. falciparum* data with T_NT_=160 days and I_0_=2100. Red: chain #1, Blue: chain #2, Green: chain #3.

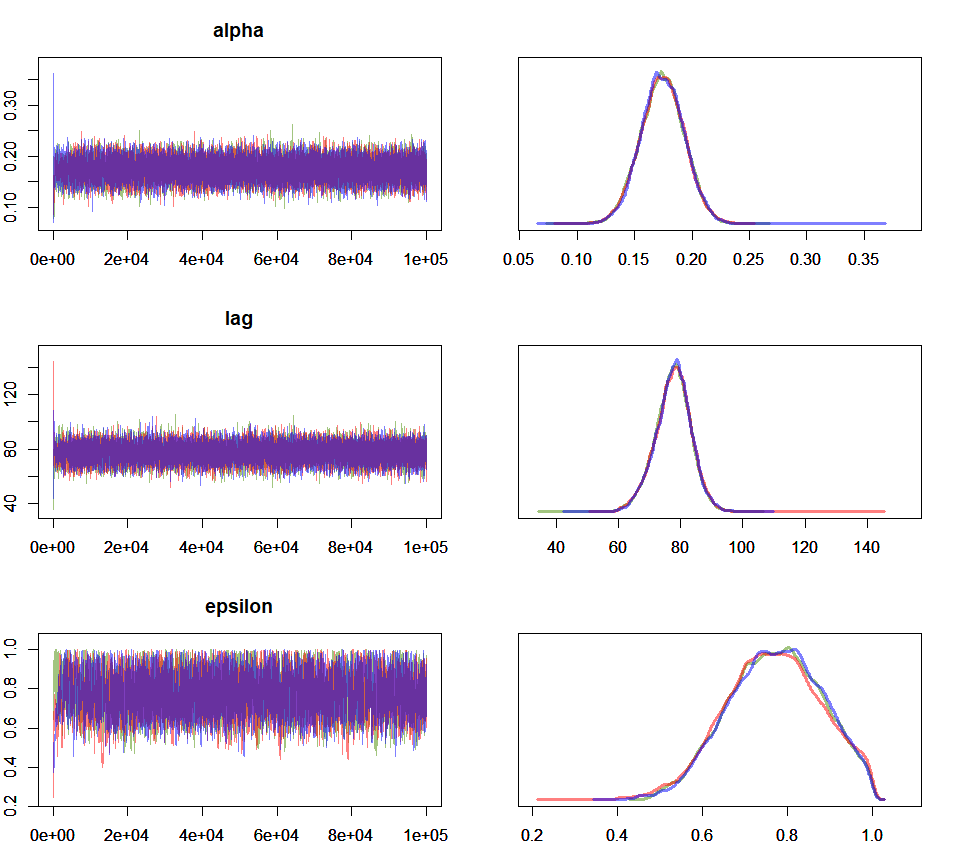

Figure 7: Trace and density plots for parameters alpha, lag and epsilon. Model fitted to *P. falciparum* data with T_NT_=160 days and I_0_=2100. Red: chain #1, Blue: chain #2, Green: chain #3.

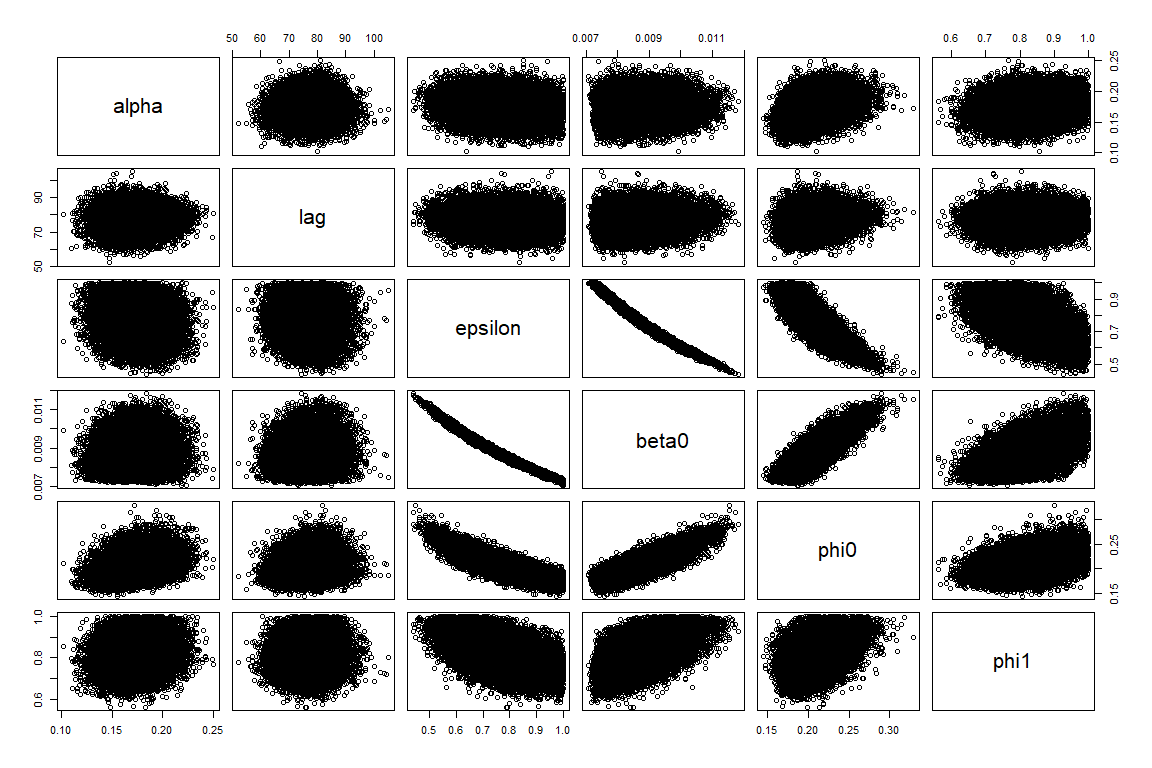

Figure 8: Correlation plots, model fitted to *P. falciparum* data with T_NT_=160 days and I_0_=2100.

##### 3.3.4 *P. vivax* model

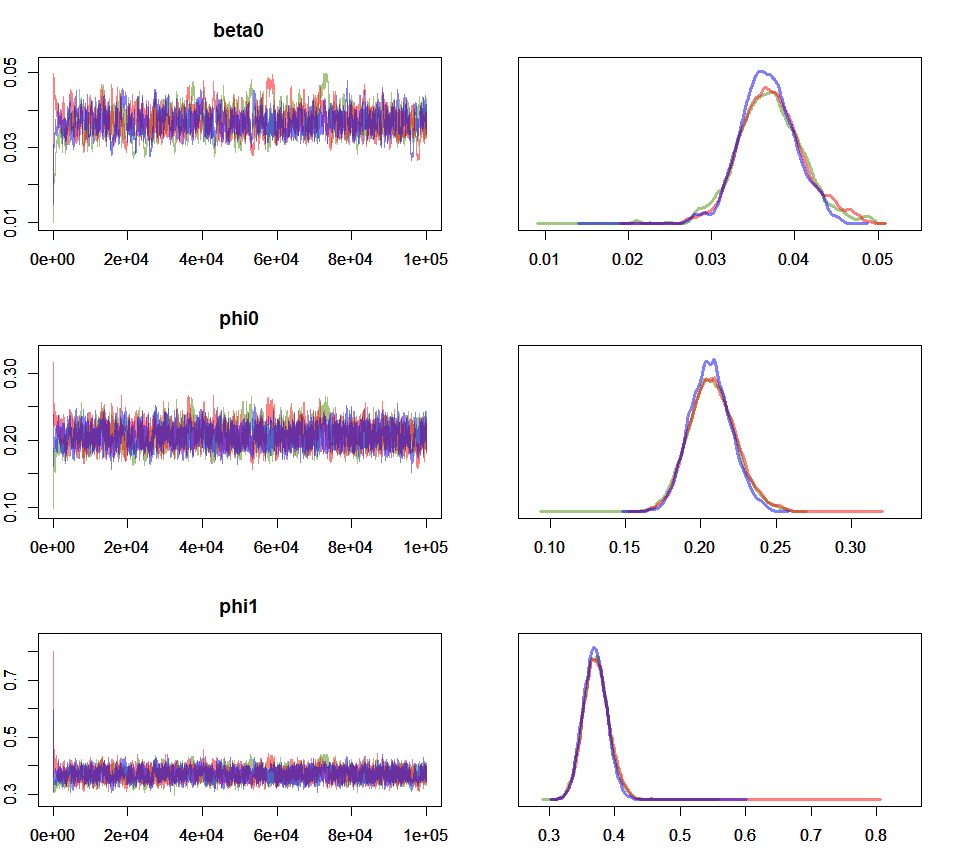

Figure 9: Trace and density plots for parameters beta0, phi0 and phi1. Model fitted to *P. vivax* data with T_NT_=70 days and I_0_=1650. Red: chain #1, Blue: chain #2, Green: chain #3.

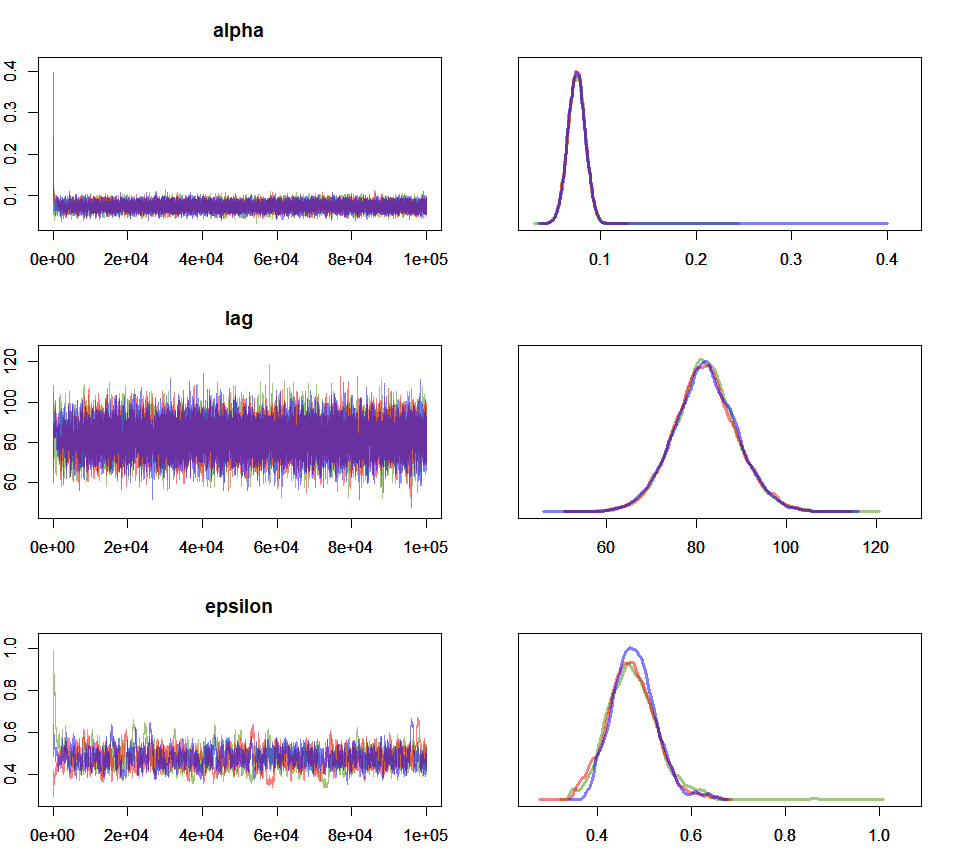

Figure 10: Trace and density plots for parameters alpha, lag and epsilon. Model fitted to *P. vivax* data with T_NT_=70 days and I_0_=1650. Red: chain #1, Blue: chain #2, Green: chain #3.

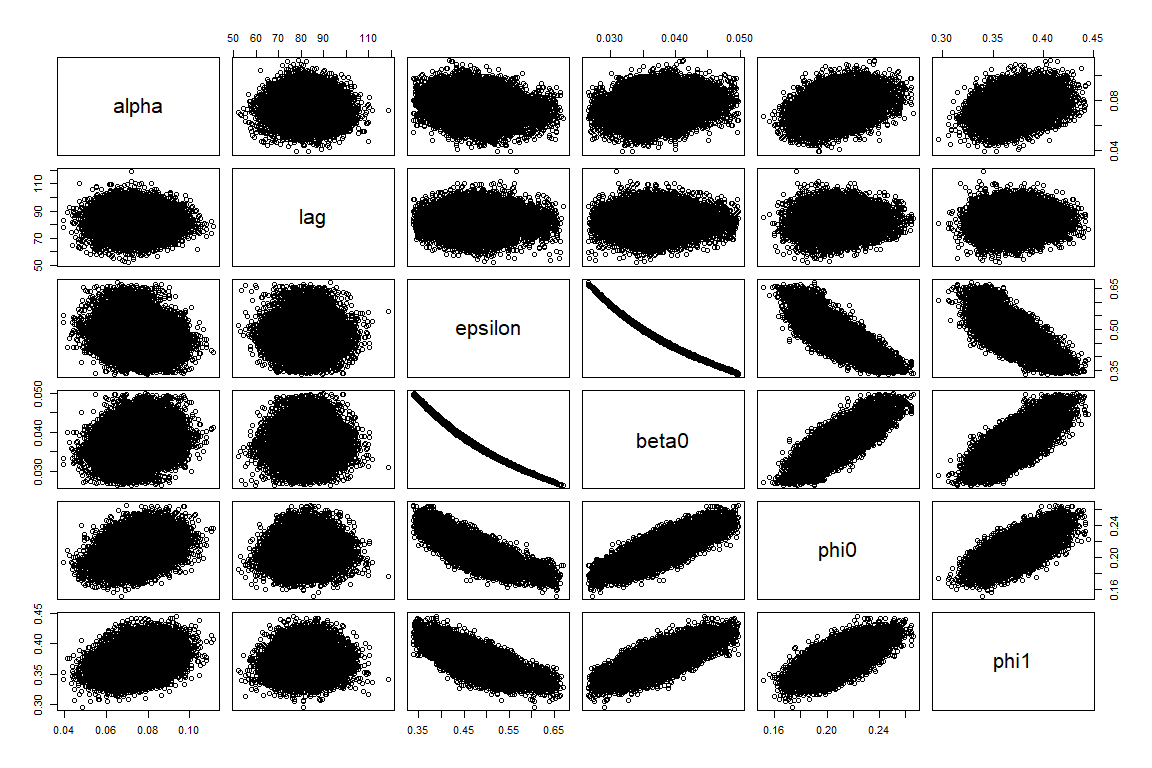

Figure 11: Correlation plots, model fitted to *P. vivax* data with T_NT_=70 days and I_0_=1650.
